# Covid-19 in the California State Prison System: An Observational Study of Decarceration, Ongoing Risks, and Risk Factors

**DOI:** 10.1101/2021.03.04.21252942

**Authors:** Elizabeth T. Chin, Theresa Ryckman, Lea Prince, David Leidner, Fernando Alarid-Escudero, Jason R. Andrews, Joshua A. Salomon, David M. Studdert, Jeremy D. Goldhaber-Fiebert

## Abstract

**Background:** Correctional institutions nationwide are seeking to mitigate Covid-19-related risks.

**Objective:** To quantify changes to California’s prison population since the pandemic began and identify risk factors for Covid-19 infection.

**Design:** We described residents’ demographic characteristics, health status, Covid-19 risk scores, room occupancy, and labor participation. We used Cox proportional hazard models to estimate the association between rates of Covid-19 infection and room occupancy and out-of-room labor, respectively.

**Setting:** California state prisons (March 1-October 10, 2020).

**Participants:** Residents of California state prisons.

**Measurements:** Changes in the incarcerated population’s size, composition, housing, and activities. For the risk factor analysis, the exposure variables were room type (cells vs dormitories) and labor participation (any room occupant participating in the prior 2 weeks) and the outcome variable was incident Covid-19 case rates.

**Results:** The incarcerated population decreased 19.1% (119,401 to 96,623) during the study period.On October 10, 2020, 11.5% of residents were aged ≥60, 18.3% had high Covid-19 risk scores, 31.0% participated in out-of-room labor, and 14.8% lived in rooms with ≥10 occupants. Nearly 40% of residents with high Covid-19 risk scores lived in dormitories. In 9 prisons with major outbreaks (6,928 rooms; 21,750 residents), dormitory residents had higher infection rates than cell residents (adjusted hazard ratio [AHR], 2.51 95%CI, 2.25-2.80) and residents of rooms with labor participation had higher rates than residents of other rooms (AHR, 1.56; 95%CI, 1.39-1.74).

**Limitations:** Inability to measure density of residents’ living conditions or contact networks among residents and staff.

**Conclusion:** Despite reductions in room occupancy and mixing, California prisons still house many medically vulnerable residents in risky settings. Reducing risks further requires a combination of strategies, including rehousing, decarceration, and vaccination.

**Funding Sources:** Horowitz Family Foundation; National Institute on Drug Abuse; National Science Foundation Graduate Research Fellowship; Open Society Foundations.

## Introduction

Over 380,000 incarcerated people in the United States were diagnosed with severe acute respiratory syndrome coronavirus 2 through February 2021, and approximately 2,300 died (1). Prisons are vulnerable to rapid viral spread, given their population density and the infeasibility of standard distancing measures (2–8). Covid-19-related health outcomes appear worse among incarcerated people than in the general population (3,9,10).

Correctional systems face difficult trade-offs in their attempts to control Covid-19 transmission. Early releases reduce crowding but may cause public unrest (11). Curtailing in-prison activities (e.g., work or group therapy) limits mixing but is disruptive and may have adverse health effects (12–13). The federal government has issued guidance (14–15) on measures to reduce Covid-19 transmission in correctional settings, but the recommendations lack specificity and evidence of efficacy. The evidence base in this area remains extremely limited.

The California Department of Corrections and Rehabilitation (CDCR) manages the country’s second largest prison system, with 35 institutions that housed ~120,000 residents in early 2020. CDCR has undertaken multiple interventions to prevent and contain Covid-19 outbreaks. This study describes how CDCR’s incarcerated population has changed during the pandemic, focusing on room occupancy and participation in out-of-room activities. For prisons that have experienced outbreaks, we estimate the extent to which those two factors are associated with Covid-19 infection rates.

## Methods

### IRB Approval

Stanford’s institutional review board granted approval (IRB-55835).

### Data and Variables

CDCR provided data on all incarcerated people aged ≥18 years residing in its prisons during the period March 1 through October 10, 2020. The data were provided at the person-day-level, which allowed daily tracking of changes in any time-varying information until residents’ release or death.

The data included variables indicating residents’ demographic (sex, age, and race or ethnicity) and health characteristics; location; participation in prison labor, education, and other activities; and Covid-19 testing history. The data contained each resident’s security level (1 [lowest] to 4 [highest]) which determines housing locations and eligibility for work and other activities (16). Locational information specified the room in which each resident spent the night. Rooms were defined as discrete spaces, at least partially enclosed by solid walls, and were classified as cells or dormitories of varying sizes.

The health information included indicators for diagnosed medical conditions and Covid-19 risk score. This score, developed by CDCR, is an integer-based estimate of each resident’s probability of severe health outcomes following Covid-19 infection. Scores correspond to the presence of demographic and clinical characteristics identified in the literature as risk factors for severe Covid-19-related illness, and CDCR considers scores ≥3 to indicate high-risk (Table S1).

Testing information included dates and results. CDCR has been testing residents using real-time PCR and antigen tests since April 2020. Testing expanded during the study period, eventually employing both reactive mass testing and periodic surveillance testing. Prisons experiencing large outbreaks tested residents at particularly high rates (Figure S1). By Fall 2020, all prisons were testing 5-25% of residents every two weeks.

We created variables to describe risk factors for Covid-19 exposure and transmission, based on residents’ housing situation and their participation in out-of-room activities. With respect to housing, we calculated the daily number of residents housed in each room (square footage of rooms was unavailable). This count variable had a bimodal distribution, with many residents living alone or with one other resident, or in substantially larger rooms (≥10 residents), and relatively few in between. For some analyses we dichotomized this variable, distinguishing residents in “cells” (1-2 occupants) and “dormitories” (≥3 occupants); this aligned with CDCR’s conception of the major division in room sizes.

With respect to activities, the resident-day-level data included information on residents’ out-of-room participation in labor (e.g., janitorial), education (e.g., high school classes), and other activities (e.g., religious services) (17–18). From April 2020, educational activities were confined to residents’ rooms. Consequently, we focused on labor and other out-of-room non-educational activities excluding recreation and meals which were not comprehensively tracked. We specified variables indicating whether each resident participated in each activity type and variables indicating whether each resident or a roommate did so in the previous two weeks.

Finally, we classified prisons into 5 categories based on the predominant resident security levels, housing configurations, and CDCR advice: reception centers, medical prisons, low security and general population prisons, high security prisons, and mixed security and medium security prisons (details provided in Table S2).

### Analysis

We calculated changes over the study period in the size and composition of the incarcerated population, in housing, and in participation in out-of-room activities (samples for each analysis shown in Figure S2).

We used person-level survival analysis to estimate the association between room occupancy and labor activities, respectively, and rates of Covid-19 infection. We focused on sustained within-prison transmission, therefore limiting the analysis to prisons with outbreaks involving substantial resident-to-resident spread. We defined prisons with outbreaks as those having ≥50 cumulative cases during the study period and ≥10 incident cases detected on at least one day in that period. We defined an outbreak’s start date as 14 days prior to the first day with ≥10 incident cases.

We specified several additional prison-level and resident-level eligibility criteria for the survival analysis (Figure S3). Briefly, to allow ≥90 days follow-up, we excluded prisons (n=7) with outbreaks that began after July 12, 2020; to minimize confounding, we excluded one prison with testing rates that differed substantially between cells and dormitories (Figure S4); and we excluded prisons (n=3) whose outbreaks were seeded by mass introduction of cases (e.g., San Quentin), because their epidemic growth may have been atypical. Within eligible prisons, we included residents present on the day the outbreak began who were tested for Covid-19 at least once during the 90-day period.

The observation period for eligible residents’ observation ran from the start of their prison’s outbreak until the sample collection date of their first positive test or their last negative test. Release or transfer to another prison were also censoring events.

We fit a multivariable Cox proportional hazard regression model to estimate the associations of interest. The outcome variable indicated the sample collection date for the first positive Covid-19 test result among residents who had a positive test. The main exposure variables were room occupancy at the outbreak start and room-level labor participation during the 14-day period prior to the outbreak start. We also included prison fixed effects. We assessed the appropriateness of the proportional hazards assumption by inspecting plots of Schoenfeld residuals.

We conducted sensitivity analyses. Because there may be systematic differences in how residents with higher Covid-19 risk scores or higher security levels mix with other residents, we added baseline values of these covariates. We varied the required follow-up period for prison inclusion. We allowed the observation period for residents who did not test positive, exit the prison, or die to extend to the end of the study period, regardless of their last negative test date. We estimated the model clustering standard errors at the room and prison levels.

## Results

From March 1 to October 10, 2020, the resident population of California prisons decreased from 119,401 to 96,623 (Figure 1), a reduction of 19.1% that reversed prior trends (19). High security prisons (7.0%) and medical prisons (14.4%) had the smallest relative reductions.

**Figure 1:**
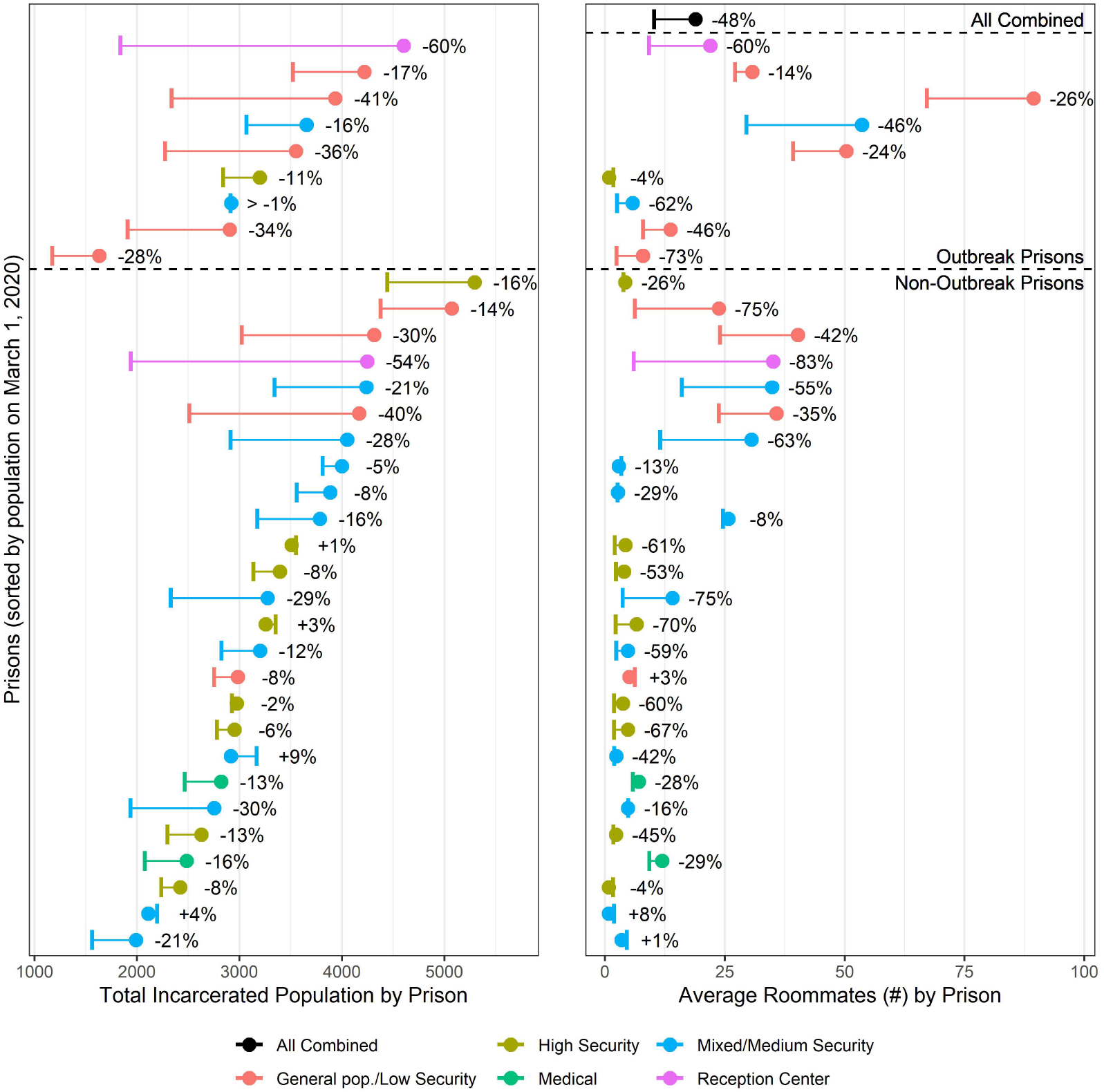
CDCR incarcerated population size and average room occupancy over time: Graph shows the change in total prison population size (left) and change in the average number of roommates an individual in that prison has (right), for each of the 35 prisons, color-coded by prison type, and for all prisons combined in black (only right panel), from March 1, 2020 (filled circle) to October 10, 2020 (vertical bar). The outbreak prisons used in the multivariate risk analysis are shown above the dashed line, and remaining prisons are shown below the dashed line.

On October 10, 2020, 96.7% of residents were male and 11.5% were aged 60 years or older (Table 1). Nearly three-quarters were Hispanic (44.5%) or non-Hispanic Black (29.5%). Forty percent had at least one medical condition, and 18.3% had a Covid-19 risk score of ≥3. The average number of residents per room decreased from 20 to 10. However, seven prisons still averaged ≥20 occupants per room in October 2020, and 25 had ≥1 room with ≥20 occupants (Figures S5-S6). The proportion of residents in dormitories decreased from 37.1% to 30.9%, though a substantial fraction of higher risk residents were still housed in dormitories (Figure S7).

**Table 1.**
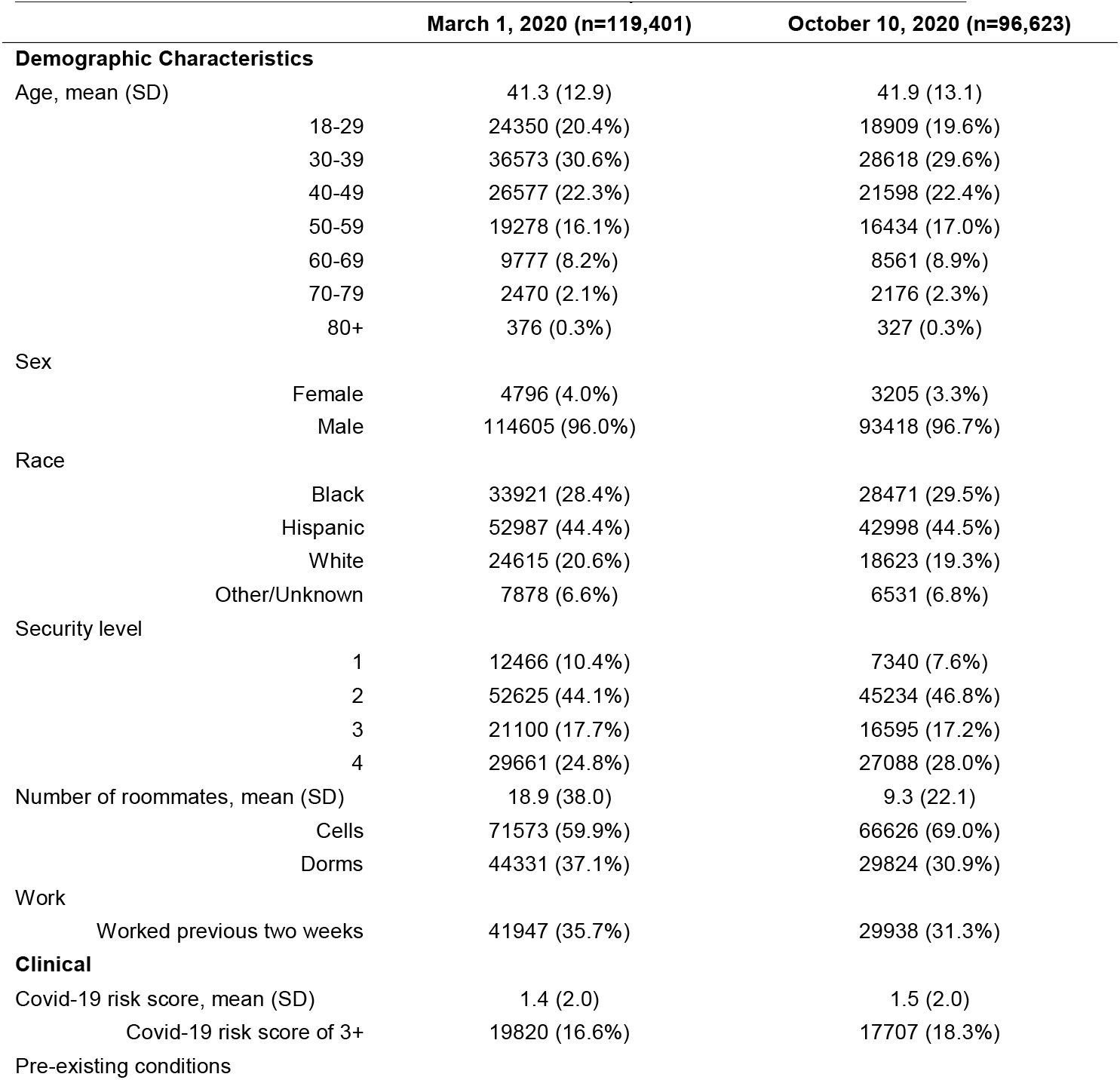
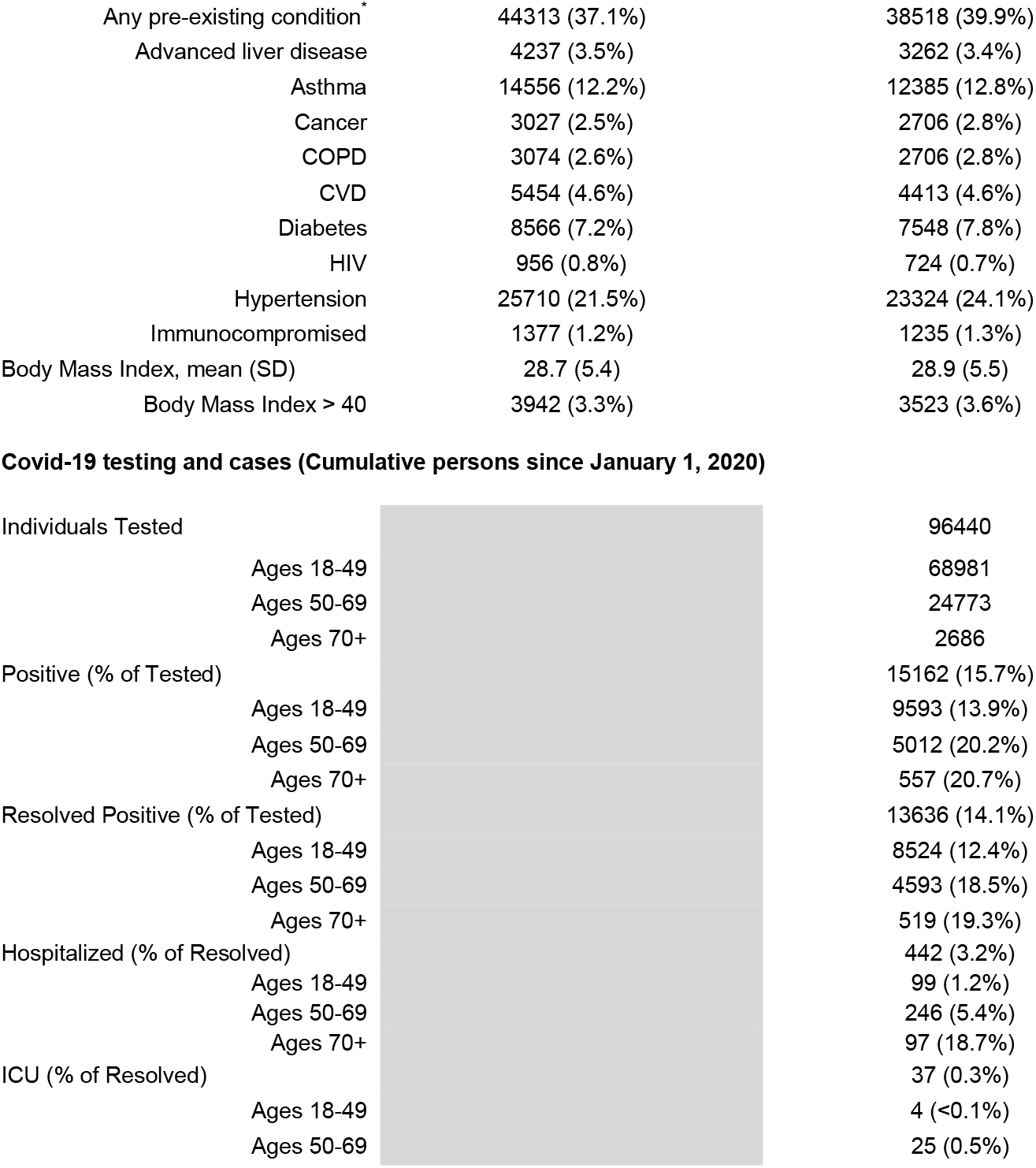
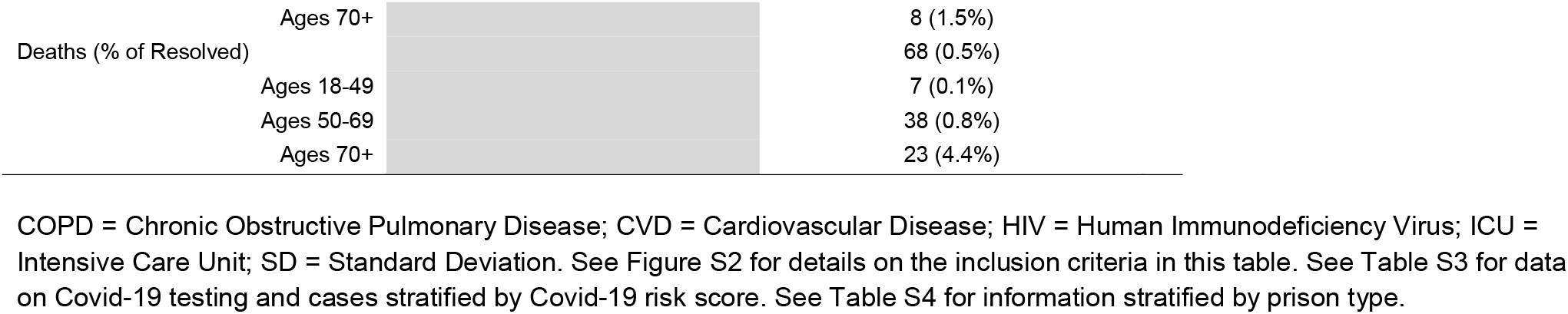
Characteristics of California State Prison Population on October 10, 2020

By October 10, 81.8% (79,046) of still-incarcerated individuals had been tested for Covid-19 at least once, and a total of 96,440 residents including those released between March and October had been tested. Of the 96,440 residents, 15,162 were positive; 13,636 of these cases resolved by October 10—3.2% were hospitalized, 0.3% were in intensive care units, and 0.5% died. Severe Covid-19 outcomes were most likely for older and higher risk residents (Table 1, Table S3).

Participation in out-of-room activities other than labor decreased precipitously between March and April 2020 and remained low through October. However, labor participation decreased only slightly during the study period (Figure 2). The lowest levels of and the largest decreases in labor participation were among residents aged ≥80 years, but more than 10% of residents in this age group and more than 20% of 70-79 year-olds were still participating in labor in October. These labor participation results were generally consistent according to resident risk measures, prison types, and outbreak history (Figures S8-S14 and Table S5).

**Figure 2:**
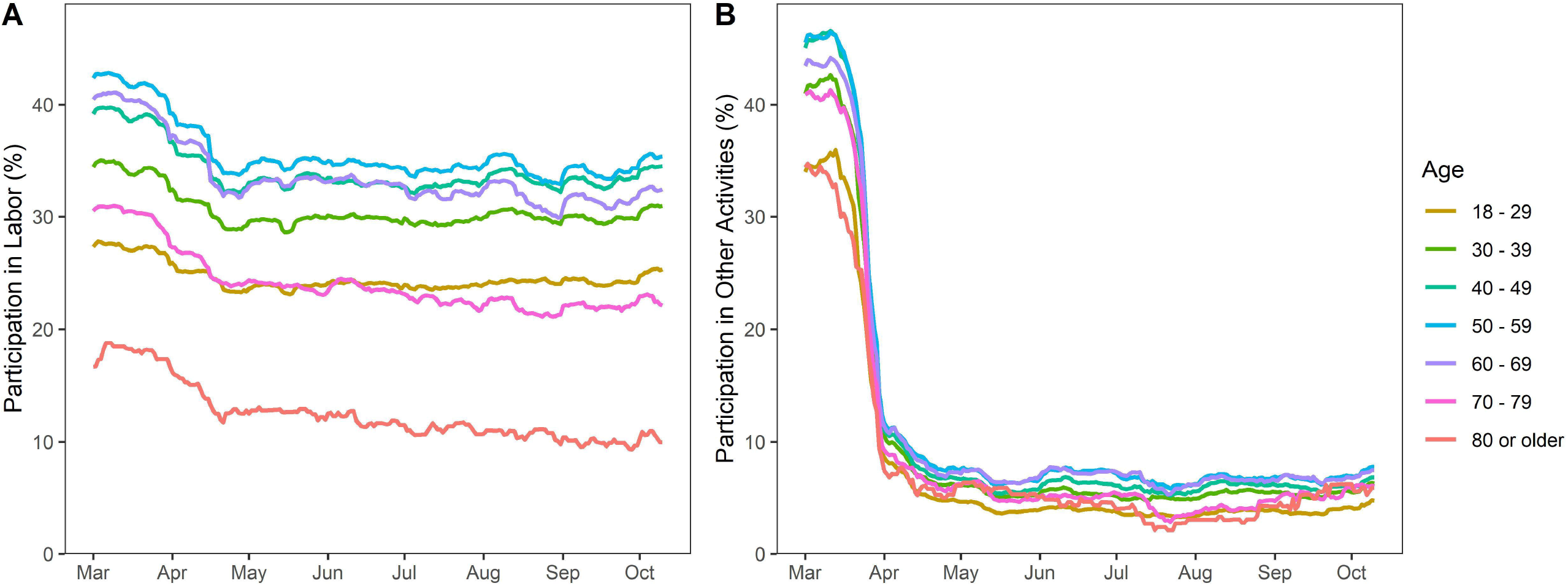
Biweekly participation in labor and other out-of-room activities by age: Graph shows rolling average participation in activities, defined as whether an individual participated in labor or other activities with at least one other person during any day in the past 2 weeks. Panels show (A) labor participation by age; (B) other participation by age. Data cover all prisons from March 1, 2020 through October 10, 2020.

In the nine prisons with outbreaks that met our eligibility criteria for the multivariable risk analysis, 21,750 eligible residents living in 6,928 rooms were tested at least once. Over 90 days, the cumulative percentage of each prison’s population tested ranged from 21.3 to 99.9%, and the cumulative percentage of each prison’s population confirmed positive ranged from 2.4 to 45.1% (Figures S15-S16).

Rates of Covid-19 infection among residents of dormitories (≥3 occupants) were more than double those among residents of cells (Adjusted hazard ratio [AHR], 2.51; 95% Confidence Interval [CI], 2.25-2.80; p<0.001) (Figure 3, Table S6). Residents of rooms with occupants participating in out-of-room labor also had higher rates of infection compared to those without participation (AHR, 1.56; 95% CI, 1.39-1.74). These differences represent a cumulative risk of infection that is 28.6 percentage points higher (95% CI, 16.7-30.6%; 62.1% vs. 33.4%) for dormitories compared to cells, and 13.1 percentage points higher (95% CI, 12.8-13.3%; 53.7% vs. 40.6%) for rooms with labor participation than for rooms without it. Estimates proved fairly robust in sensitivity analyses (Tables S6-S8; Figure S17). While, in theory, residents with higher Covid-19 risk scores may take greater precautions to avoid more severe consequences from infection, analyses that included this variable did not detect an association with lower risks.

**Figure 3.**
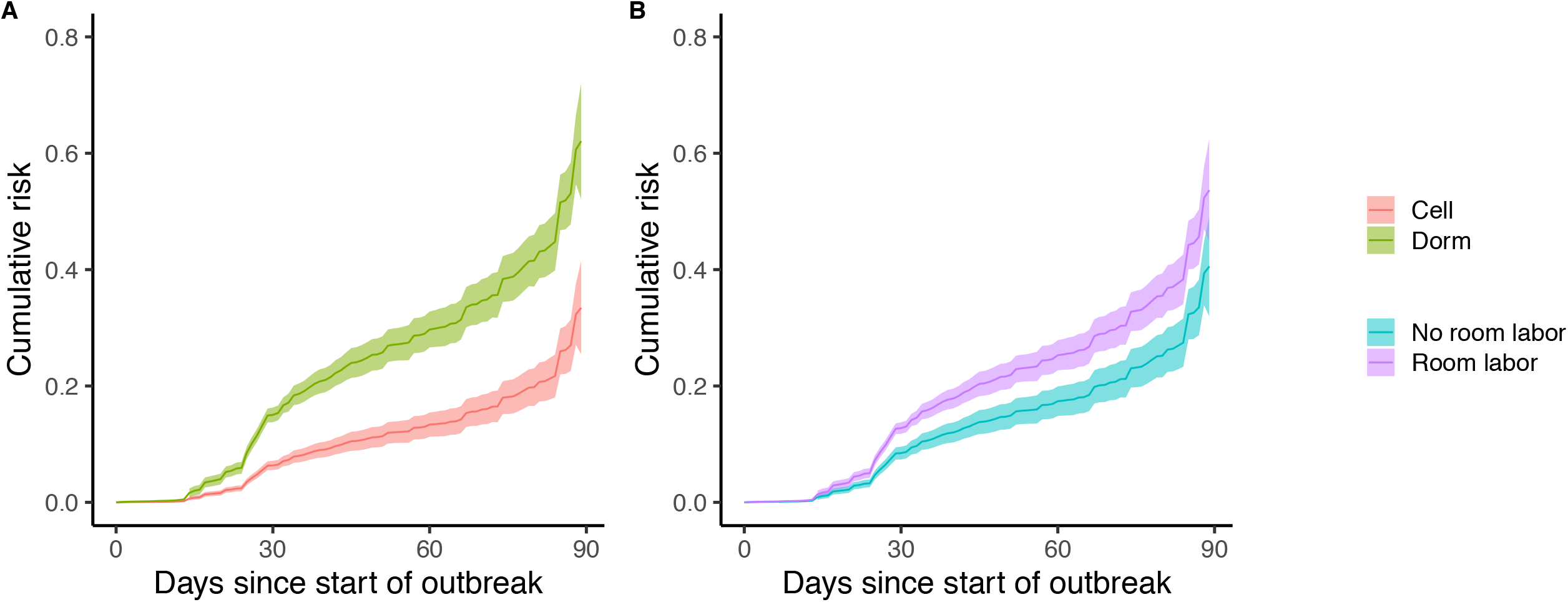
Adjusted cumulative risk of infection in outbreak prisons: Graphs show the adjusted marginal cumulative risk of infection over 90 days from the start of a prison outbreak for (A) room type and (B) room labor (i.e., whether anyone in the room participated in out-of-room labor). Data includes 90 days follow-up for 9 prisons.

## Discussion

Over a 7-month period following the onset of the Covid-19 pandemic, the resident population of California state prisons decreased by one-fifth and room occupancy halved. However, many medically vulnerable residents remained incarcerated, and a substantial proportion of them continued to live in dormitory housing and participate in work activities that involved mixing. In nine prisons that experienced large outbreaks, we found that residents living in rooms with ≥2 others had a 2.5-fold higher infection rate and that residents in a room whose occupants participated in out-of-room work had a 1.6-fold higher rate.

Since the pandemic began CDCR has taken drastic steps to mitigate transmission and stem outbreaks. Nonetheless, as of February 13, 2021, there had been 48,758 confirmed cases among residents and 205 deaths. These represent substantially higher rates of infection and mortality that state’s general population has experienced.

The patterns we observed suggest opportunities for further risk reduction. The substantial numbers of older and higher-risk residents who continue to live in high occupancy rooms is an important target group for prevention efforts. Members of those high-risk groups who participate in group activities, or who share a room with others who do, are also an important target group.

Our results are consistent with key messages from the limited evidence base on respiratory infections in incarcerated populations (2–13, 20–22). The interconnected nature of the residents of congregate institutions means that outbreaks, especially those that penetrate higher occupancy rooms, can quickly become building or prison outbreaks. Correctional facilities, nationally and internationally, should redouble efforts not only to prevent outbreaks but also to increase protective measures taken for their large, medically vulnerable subpopulations, including further reductions in the density of living arrangements (23).

Our access to person-level, daily data from a large prison system with high rates of testing created analytical opportunities that recent studies of Covid-19 in incarcerated populations have not had, including minimizing biases from selection into testing, However, our study also had limitations. First, we could not identify networks of specific contacts, a limitation that is likely to have mattered most to our estimates of the effect of labor participation. Second, we used room occupancy to indicate in-room contacts because density measures (e.g., residents per square foot) were not available. The most plausible effect of misclassifying room exposures would be to bias to the null our estimates of the effect of living in higher occupancy rooms. Third, we lacked information on some potential exposures, such as contacts with staff and during meals in common areas. Finally, although CDCR undertook extensive testing, not all residents were tested and test frequency varied across institutions. If residents of dormitories or of rooms with labor participation were tested relatively frequently, or testing there more precisely targeted infected residents, our estimates of these factors on infection risk may be biased upward.

Prisons remain particularly dangerous settings for Covid-19-related morbidity and mortality. Our study shows that thousands of vulnerable incarcerated people continue to be housed in settings where their risk of Covid-19 infection is high. Protective measures such as decarceration, testing, vaccination, and efforts to enhance vaccine uptake remain vital (24). Residents at greatest risk of transmission and infection, such as those living dormitories and participating in labor, should be prioritized. Furthermore, as correctional systems offer vaccination to residents, our study highlights the importance of prioritizing both those at highest risk of adverse outcomes following infection and those most likely to contract and spread the virus.

## Data Availability

Data set: Not available. Contact the California Department of Corrections and Rehabilitation regarding requests for data and data use agreements.

## Acknowledgements

We thank John Dunlap, Heidi Bauer and the other staff members at California Department of Corrections and Rehabilitation for providing data and assistance with interpretation of study results. We also acknowledge help from other members of the SC-COSMO consortium.

