## Supplementary Tables, Figures, and Information for "Covid-19 in the California State Prison System: An Observational Study of Decarceration, Ongoing Risks, and Risk Factors"

Table S1: Pre-Existing Medical Conditions and Covid-19 risk score

| Condition | Definition | Weighted Score |
|---|---|---|
| Age 65+ | Chronologic age of 65 years or above | 4 |
| Advanced liver disease | Has advanced liver disease (cirrhosis/end stage liver disease) | 2 |
| Asthma | Persistent asthma (moderate or severe) as defined by the California Correctional Health Care Services (CCHCS) asthma condition specifications | 1 |
| Cancer | High risk cancer as defined by the CCHCS cancer condition specifications (excludes most diagnoses of skin cancer and “personal history of” cancers”) | 2 |
| Chronic Lung Disease (other) | Has cystic fibrosis, pneumoconiosis, or pulmonary fibrosis | 1 |
| Chronic Obstructive Pulmonary Disease (COPD) | Has Chronic Obstructive Pulmonary Disease | 2 |
| Cardiovascular Disease (CVD) | Has any of the following:” cerebrovascular disease, congestive heart failure, congenital heart disease, ischemic heart disease, peripheral vascular disease, thromboembolic disease, and valvular disease | 1 |
| Cardiovascular Disease (CVD; high risk) | Is high risk for any of the following:” cerebrovascular disease, congestive heart failure, congenital heart disease, ischemic heart disease, peripheral vascular disease, thromboembolic disease, and valvular disease | 1 |
| Diabetes | Has diabetes | 1 |
| Diabetes (high risk) | Meets the criteria for high risk diabetes as defined by the CCHS diabetes condition specifications | 1 |
| HIV | Has HIV | 1 |
| HIV (poorly controlled) | Has HIV with a CD4 count < 200 | 1 |
| Immunocompromised | Has any of the following: aplastic anemia, histiocytosis, immunosuppressed, organ | 2 |

| | | |
|---|---|---|
| | transplant, other transplant | |
| Morbid Obesity | Body Mass Index of 40 or above | 1 |
| Other Chronic Conditions | Has any of the following with a high-risk rating: hypertension, coccidiomycosis, connective tissue disorder, dementia/Parkinson's disease, endocrine disorder, multiple sclerosis, myasthenia gravis, neurologic disorder, vasculitis | 1 |
| On Dialysis | On hemodialysis | 2 |
| Pregnant | Actively pregnant | 1 |

Note: the Covid-19 risk score is calculated as the sum of the weighted score.

We classified prisons into 5 categories, based on the predominant resident security levels, housing configurations, and CDCR advice: 1) reception centers, where new arrivals are processed; 2) medical prisons, which house people with severe chronic health conditions, physical disabilities, and mental illnesses; 3) low security and general population prisons, where housing for security levels 1-2 and dormitories predominated; 4) high security prisons, where housing for security levels 3-4 and cells predominated; and 5) mixed security and medium security prisons, which contained a mixture of security levels and housing types. Table S2 shows the categorization of each of the CDCR prisons.

Table S2: Prison Types

| **Prison** | **County** | **Prison category** |
|---|---|---|
| Avenal State Prison | Kings | General population / Low Security |
| California City Correctional Facility | Kern | Mixed / Medium Security |
| Calipatria State Prison | Imperial | High Security |
| California Correctional Center | Lassen | General population / Low Security |
| California Correctional Institution | Kern | Mixed / Medium Security |
| Central California Women's Facility | Madera | Mixed / Medium Security |
| California State Prison, Centinela | Imperial | High Security |
| California Health Care Facility | San Joaquin | Medical |
| California Institution for Men | San Bernardino | General population / Low Security |
| California Institution for Women | Riverside | General population / Low Security |
| California Men's Colony | San Luis Obispo | Mixed / Medium Security |
| California Medical Facility | Solano | Medical |
| California State Prison, Corcoran | Kings | Mixed / Medium Security |
| California Rehabilitation Center | Riverside | General population / Low Security |
| Correctional Training Facility | Monterey | General population / Low Security |
| Chuckawalla Valley State Prison | Riverside | General population / Low Security |
| Deuel Vocational Institution | San Joaquin | Mixed / Medium Security |
| Folsom State Prison | Sacramento | Mixed / Medium Security |
| High Desert State Prison | Lassen | High Security |
| Ironwood State Prison | Riverside | Mixed / Medium Security |
| Kern Valley State Prison | Kern | High Security |
| California State Prison, Los Angeles County | Los Angeles | High Security |
| Mule Creek State Prison | Amador | Mixed / Medium Security |
| North Kern State Prison | Kern | Reception Center |
| Pelican Bay State Prison | Del Norte | High Security |

| | | |
|---|---|---|
| Pleasant Valley State Prison | Fresno | Mixed / Medium Security |
| Richard J. Donovan Correctional Facility | San Diego | Mixed / Medium Security |
| California State Prison, Sacramento | Sacramento | High Security |
| California Substance Abuse Treatment Facility and State Prison, Corcoran | Kings | High Security |
| Sierra Conservation Center | Tuolumne | General population / Low Security |
| California State Prison, Solano | Solano | Mixed / Medium Security |
| San Quentin State Prison | Marin | Mixed / Medium Security |
| Salinas Valley State Prison | Monterey | High Security |
| Valley State Prison | Madera | General population / Low Security |
| Wasco State Prison | Kern | Reception Center |

Table S3: Covid-19 testing and cases (Cumulative persons since January 1, 2020) stratified by Covid-19 risk score

| | |
|---|---|
| Individuals Tested | 96440 |
| Covid-19 risk score < 3 | 78041 |
| Covid-19 risk score of 3+ | 18399 |
| Positive (% of Tested) | 15162 (15.7%) |
| Covid-19 risk score < 3 | 11946 (15.3%) |
| Covid-19 risk score of 3+ | 3216 (17.5%) |
| Resolved Positive (% of Tested) | 13636 (14.1%) |
| Covid-19 risk score < 3 | 10658 (13.7%) |
| Covid-19 risk score of 3+ | 2978 (16.2%) |
| Hospitalized (% of Resolved) | 442 (3.2%) |
| Covid-19 risk score < 3 | 152 (1.4%) |
| Covid-19 risk score of 3+ | 290 (9.7%) |
| ICU (% of Resolved) | 37 (0.3%) |
| Covid-19 risk score < 3 | 5 (<0.1%) |
| Covid-19 risk score of 3+ | 32 (1.1%) |
| Deaths (% of Resolved) | 68 (0.5%) |
| Covid-19 risk score < 3 | 9 (0.1%) |
| Covid-19 risk score of 3+ | 59 (2.0%) |

Table S4: Change in prison characteristics between March and October by prison type

| | March 1, 2020 | | | | | | October 10, 2020 | | | | | |
|---|---|---|---|---|---|---|---|---|---|---|---|---|
| | Overall (n=119,401) | Reception Center (n=8,851; 7.4%) | General pop / Low Security (n=32,798; 27.5%) | Mixed/Medium Security (n=42,800; 35.8%) | High Security (n=29,643; 24.8%) | Medical (n=5,309; 4.4%) | Overall (n=96,623) | Reception Center (n=3,780; 3.9%) | General pop / Low Security (n=23,894; 24.7%) | Mixed/Medium Security (n=36,823; 38.1%) | High Security (n=27,581; 28.5%) | Medical (n=4,545; 4.7%) |
| **Demographic Characteristics** | | | | | | | | | | | | |
| Age, mean (SD) | 41.3 (12.9) | 35.4 (10.6) | 42.9 (12.8) | 42.0 (13.4) | 38.6 (11.0) | 51.3 (15.3) | 41.9 (13.1) | 35.5 (10.6) | 43.8 (12.9) | 42.7 (13.6) | 38.5 (11.0) | 52.1 (15.0) |
| 18-29 | 24350 (20.4%) | 3074 (34.7%) | 5342 (16.3%) | 8766 (20.5%) | 6717 (22.7%) | 451 (8.5%) | 18909 (19.6%) | 1297 (34.3%) | 3682 (15.4%) | 7119 (19.3%) | 6466 (23.4%) | 345 (7.6%) |
| 30-39 | 36573(30.6%) | 3118 (35.2%) | 9253 (28.2%) | 12280 (28.7%) | 10959 (37.0%) | 963 (18.1%) | 28618 (29.6%) | 1348 (35.7%) | 6180 (25.9%) | 10183 (27.7%) | 10153 (36.8%) | 754 (16.6%) |
| 40-49 | 26577 (22.3%) | 1576 (17.8%) | 7922 (24.2%) | 9410 (22.0%) | 6706 (22.6%) | 963 (18.1%) | 21598 (22.4%) | 684 (18.1%) | 5848 (24.5%) | 8077 (21.9%) | 6159 (22.3%) | 830 (18.3%) |
| 50-59 | 19278 (16.1%) | 816 (9.2%) | 6418 (19.6%) | 7116 (16.6%) | 3746 (12.6%) | 1182 (22.3%) | 16434 (17%) | 340 (9.0%) | 5115 (21.4%) | 6502 (17.7%) | 3429 (12.4%) | 1048 (23.1%) |
| 60-69 | 9777 (8.2%) | 241 (2.7%) | 3172 (9.7%) | 3936 (9.2%) | 1339 (4.5%) | 1089 (20.5%) | 8561 (8.9%) | 93 (2.5%) | 2565 (10.7%) | 3717 (10.1%) | 1209 (4.4%) | 977 (21.5%) |
| 70-79 | 2470 (2.1%) | 23 (0.3%) | 620 (1.9%) | 1163 (2.7%) | 150 (0.5%) | 514 (9.7%) | 2176 (2.3%) | 15 (0.4%) | 449 (1.9%) | 1106 (3.0%) | 138 (0.5%) | 468 (10.3%) |
| 80+ | 376 (0.3%) | 3 (0.0%) | 71 (0.2%) | 129 (0.3%) | 26 (0.1%) | 147 (2.8%) | 327 (0.3%) | 3 (0.1%) | 55 (0.2%) | 119 (0.3%) | 27 (0.1%) | 123 (2.7%) |
| Sex | | | | | | | | | | | | |
| Women | 4796 (4.0%) | 0 (0.0%) | 1634 (5.0%) | 3162 (7.4%) | 0 (0.0%) | 0 (0.0%) | 3205 (3.3%) | 0 (0.0%) | 1174 (4.9%) | 2031 (5.5%) | 0 (0.0%) | 0 (0.0%) |
| Men | 114605 (96.0%) | 8851 (100.0%) | 31164 (95.0%) | 39638 (92.6%) | 29643 (100.0%) | 5309 (100.0%) | 93418 (96.7%) | 3780 (100.0%) | 22720 (95.1%) | 34792 (94.5%) | 27581 (100.0%) | 4545 (100.0%) |
| Race | | | | | | | | | | | | |
| Black | 33921 (28.4%) | 2236 (25.3%) | 7113 (21.7%) | 12633 (29.5%) | 9839 (33.2%) | 2100 (39.6%) | 28471 (29.5%) | 1146 (30.3%) | 4985 (20.9%) | 11168 (30.3%) | 9340 (33.9%) | 1832 (40.3%) |
| Hispanic | 52987 (44.4%) | 4856 (54.9%) | 14955 (45.6%) | 17820 (41.6%) | 14139 (47.7%) | 1217 (22.9%) | 42998 (44.5%) | 1843 (48.8%) | 11306 (47.3%) | 15582 (42.3%) | 13250 (48.0%) | 1017 (22.4%) |
| White | 24615 (20.6%) | 1431 (16.2%) | 8325 (25.4%) | 9189 (21.5%) | 4106 (13.9%) | 1564 (29.5%) | 18623 (19.3%) | 617 (16.3%) | 5761 (24.1%) | 7377 (20.0%) | 3554 (12.9%) | 1314 (28.9%) |

| | | | | | | | | | | | | |
|---|---|---|---|---|---|---|---|---|---|---|---|---|
| Other/Unknown | 7878 (6.6%) | 328 (3.7%) | 2405 (7.3%) | 3158 (7.4%) | 1559 (5.3%) | 428 (8.1%) | 6521 (6.8%) | 174 (4.6%) | 1842 (7.7%) | 2696 (7.3%) | 1437 (5.2%) | 382 (8.4%) |
| Security level | | | | | | | | | | | | |
| 1 | 12466 (10.4%) | 1399 (15.8%) | 6220 (19.0%) | 3403 (8.0%) | 1098 (3.7%) | 346 (6.5%) | 7340 (7.6%) | 531 (14.0%) | 3970 (16.6%) | 2139 (5.8%) | 493 (1.8%) | 207 (4.6%) |
| 2 | 52625 (44.1%) | 2410 (27.2%) | 23671 (72.2%) | 18418 (43.0%) | 4812 (16.2%) | 3314 (62.4%) | 45234 (46.8%) | 1241 (32.8%) | 18237 (76.3%) | 18684 (50.7%) | 4081 (14.8%) | 2991 (65.8%) |
| 3 | 21100 (17.7%) | 2184 (24.7%) | 2233 (6.8%) | 11523 (26.9%) | 4487 (15.1%) | 673 (12.7%) | 16595 (17.2%) | 1379 (36.5%) | 1277 (5.3%) | 8758 (23.8%) | 4675 (17.0%) | 506 (11.1%) |
| 4 | 29661 (24.8%) | 491 (5.5%) | 471 (1.4%) | 8517 (19.9%) | 19225 (64.9%) | 957 (18.0%) | 27088 (28.0%) | 347 (9.2%) | 365 (1.5%) | 7224 (19.6%) | 18317 (66.4%) | 835 (18.4%) |
| Number of roommates, mean (SD) | 18.9 (38.0) | 30.4 (44.6) | 35.6 (43.1) | 16.3 (40.2) | 3.7 (14.3) | 9.3 (20.9) | 9.3 (22.1) | 6.6 (13.7) | 21.6 (31.3) | 7.9 (21.1) | 1.3 (2.6) | 6.4 (15.3) |
| Cells | 71573 (59.9%) | 4095 (46.3%) | 9137 (27.9%) | 29428 (68.8%) | 25562 (86.2%) | 3351 (63.1%) | 66626 (69.0%) | 2953 (78.1%) | 7972 (33.4%) | 27528 (74.8%) | 25077 (90.9%) | 3096 (68.1%) |
| Dorms | 44331 (37.1%) | 2742 (31.0%) | 23178 (70.7%) | 12733 (29.8%) | 3892 (13.1%) | 1786 (33.6%) | 29824 (30.9%) | 789 (20.9%) | 15917 (66.6%) | 9186 (24.9%) | 2489 (9.2%) | 1443 (31.7%) |
| Work | | | | | | | | | | | | |
| Worked previous two weeks | 41947 (35.7%) | 915 (11.5%) | 14625 (44.9%) | 15361 (36.3%) | 9858 (33.4%) | 1188 (22.7%) | 29938 (31.3%) | 763 (22.9%) | 9339 (39.2%) | 10693 (29.2%) | 8345 (30.4%) | 798 (17.7%) |
| **Clinical** | | | | | | | | | | | | |
| CDCR Covid-19 risk score, mean (SD) | 1.4 (2.0) | 0.7 (1.0) | 1.3 (1.8) | 1.6 (2.1) | 1.0 (1.4) | 3.4 (3.4) | 1.5 (2.0) | 0.9 (1.1) | 1.4 (1.8) | 1.7 (2.2) | 1.1 (1.4) | 3.5 (3.3) |
| 3+ | 19820 (16.6%) | 452 (5.1%) | 5139 (15.7%) | 8493 (19.8%) | 3276 (11.1%) | 2460 (46.3%) | 17707 (18.3%) | 264 (7.0%) | 4061 (17.0%) | 7968 (21.6%) | 3180 (11.5%) | 2234 (49.2%) |
| Pre-existing conditions | | | | | | | | | | | | |
| Any pre-existing condition* | 44314 (37.1%) | 2088 (23.6%) | 11301 (34.5%) | 17323 (40.5%) | 10070 (34.0%) | 3531 (66.5%) | 38306 (39.6%) | 1039 (27.5%) | 8841 (37.0%) | 15861 (43.1%) | 9636 (34.9%) | 3141 (69.1%) |
| Advanced liver disease | 4237 (3.5%) | 56 (0.6%) | 840 (2.6%) | 1953 (4.6%) | 843 (2.8%) | 545 (10.3%) | 3262 (3.4%) | 31 (0.8%) | 529 (2.2%) | 1585 (4.3%) | 670 (2.4%) | 447 (9.8%) |

| | | | | | | | | | | | | |
|---|---|---|---|---|---|---|---|---|---|---|---|---|
| Asthma | 14556 (12.2%) | 977 (11.0%) | 3120 (9.5%) | 5507 (12.9%) | 4111 (13.9%) | 841 (15.8%) | 12385 (12.8%) | 473 (12.5%) | 2310 (9.7%) | 4910 (13.3%) | 3943 (14.3%) | 749 (16.5%) |
| Cancer | 3027 (2.5%) | 60 (0.7%) | 747 (2.3%) | 1423 (3.3%) | 376 (1.3%) | 421 (7.9%) | 2706 (2.8%) | 35 (0.9%) | 604 (2.5%) | 1304 (3.5%) | 386 (1.4%) | 377 (8.3%) |
| COPD | 3074 (2.6%) | 35 (0.4%) | 731 (2.2%) | 1274 (3.0%) | 392 (1.3%) | 642 (12.1%) | 2706 (2.8%) | 21 (0.6%) | 560 (2.3%) | 1205 (3.3%) | 361 (1.3%) | 559 (12.3%) |
| CVD | 5454 (4.6%) | 99 (1.1%) | 1090 (3.3%) | 2364 (5.5%) | 936 (3.2%) | 965 (18.2%) | 4413 (4.6%) | 51 (1.3%) | 762 (3.2%) | 1939 (5.3%) | 858 (3.1%) | 803 (17.7%) |
| Diabetes | 8566 (7.2%) | 305 (3.4%) | 2461 (7.5%) | 3355 (7.8%) | 1341 (4.5%) | 1104 (20.8%) | 7548 (7.8%) | 145 (3.8%) | 1927 (8.1%) | 3198 (8.7%) | 1280 (4.6%) | 998 (22.0%) |
| HIV | 956 (0.8%) | 42 (0.5%) | 226 (0.7%) | 439 (1.0%) | 105 (0.4%) | 144 (2.7%) | 724 (0.7%) | 24 (0.6%) | 123 (0.5%) | 368 (1.0%) | 94 (0.3%) | 115 (2.5%) |
| Hypertension | 25710 (21.5%) | 979 (11.1%) | 6821 (20.8%) | 10089 (23.6%) | 5352 (18.1%) | 2469 (46.5%) | 23324 (24.1%) | 504 (13.3%) | 5631 (23.6%) | 9696 (26.3%) | 5251 (19.0%) | 2242 (49.3%) |
| Immunocompromised | 1377 (1.2%) | 13 (0.1%) | 278 (0.8%) | 622 (1.5%) | 244 (0.8%) | 220 (4.1%) | 1235 (1.3%) | 9 (0.2%) | 203 (0.8%) | 575 (1.6%) | 248 (0.9%) | 200 (4.4%) |
| Body Mass Index, mean (SD) | 28.7 (5.4) | 28.4 (5) | 29.1 (5.4) | 28.8 (5.4) | 28.1 (5) | 28.7 (6.8) | 28.9 (5.5) | 29.1 (5.1) | 29.3 (5.5) | 29.1 (5.5) | 28.4 (5.2) | 29.2 (7) |
| Body Mass Index > 40 | 3942 (3.3%) | 261 (2.9%) | 1169 (3.6%) | 1572 (3.7%) | 697 (2.4%) | 243 (4.6%) | 3523 (3.6%) | 144 (3.8%) | 911 (3.8%) | 1475 (4.0%) | 752 (2.7%) | 241 (5.3%) |
| **Covid-19 testing and cases (Cumulative persons since January 1, 2020)** | | | | | | | | | | | | |
| Individuals Tested | | | | | | | n=96440 | n=4480 | n=27920 | n=37490 | n=21727 | n=4823 |
| Positive (% of Tested) | | | | | | | 15162 (15.7%) | 289 (6.5%) | 8749 (31.3%) | 5284 (14.1%) | 813 (3.7%) | 27 (0.6%) |
| Positive and Resolved (% of Tested) | | | | | | | 1363 (14.1%) | 245 (5.5%) | 7635 (27.3%) | 4997 (13.3%) | 732 (3.4%) | 27 (0.6%) |
| Hospitalized (% of Resolved) | | | | | | | 442 (3.2%) | 2 (0.8%) | 252 (3.3%) | 166 (3.3%) | 21 (2.9%) | 1 (3.7%) |
| ICU (% of Resolved) | | | | | | | 37 (0.3%) | 0 (0.0%) | 12 (0.2%) | 25 (0.5%) | 0 (0.0%) | 0 (0.0%) |
| Deaths (% of Resolved) | | | | | | | 68 (0.5%) | 289 (6.5%) | 8749 (31.3%) | 5284 (14.1%) | 813 (3.7%) | 27 (0.6%) |

COPD = Chronic Obstructive Pulmonary Disease; CVD = Cardiovascular Disease; HIV = Human Immunodeficiency Virus; ICU = Intensive Care Unit; SD = Standard Deviation.

Figure S1:  Testing patterns for prisons experiencing outbreaks with >90 days

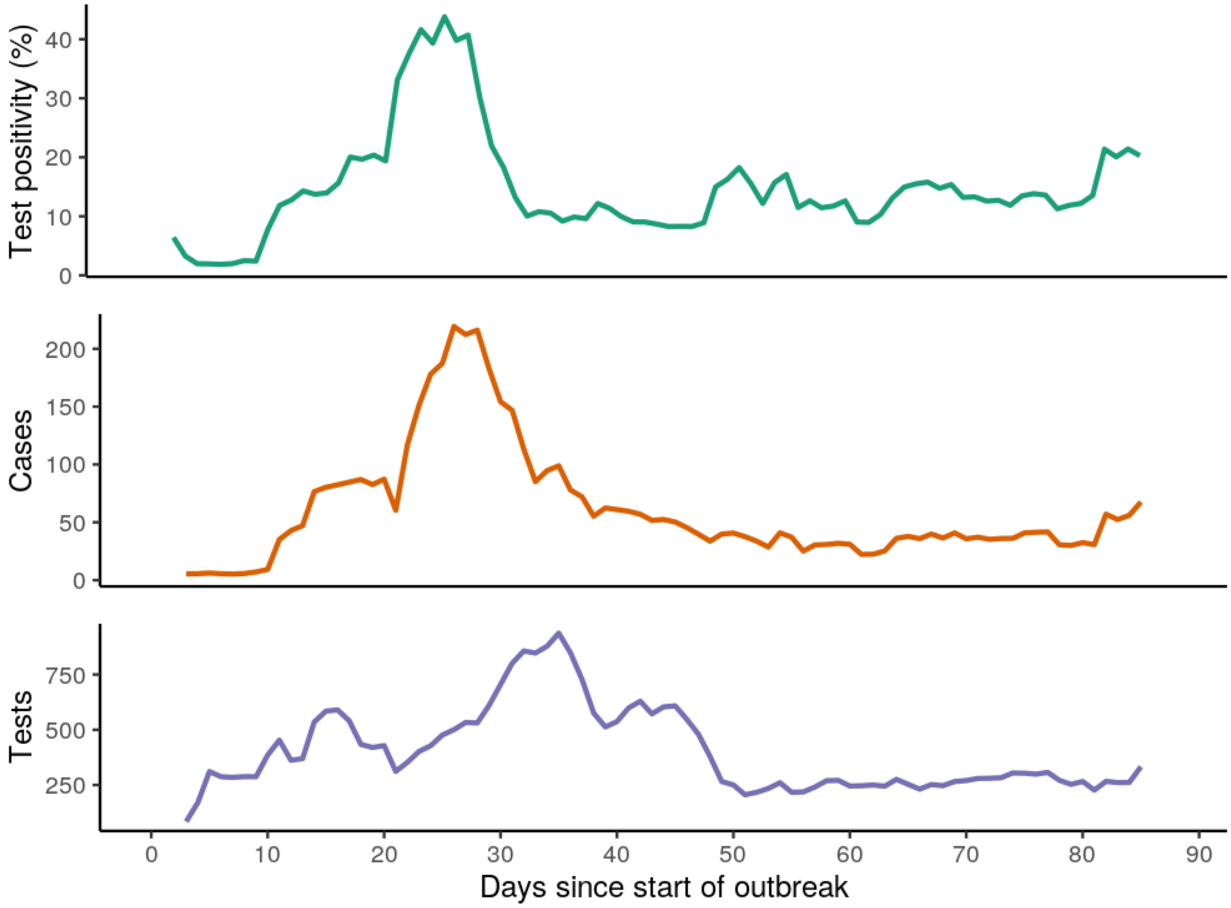


Notes: Graph shows rolling average test positivity, cases, and tests. Individuals are censored after their first positive test (when they are counted as a case). Data includes 90 days follow-up for 9 prisons.

Figure S2: Inclusion and exclusion criteria for results reported in Table 1, Figure 1, and Figure 2

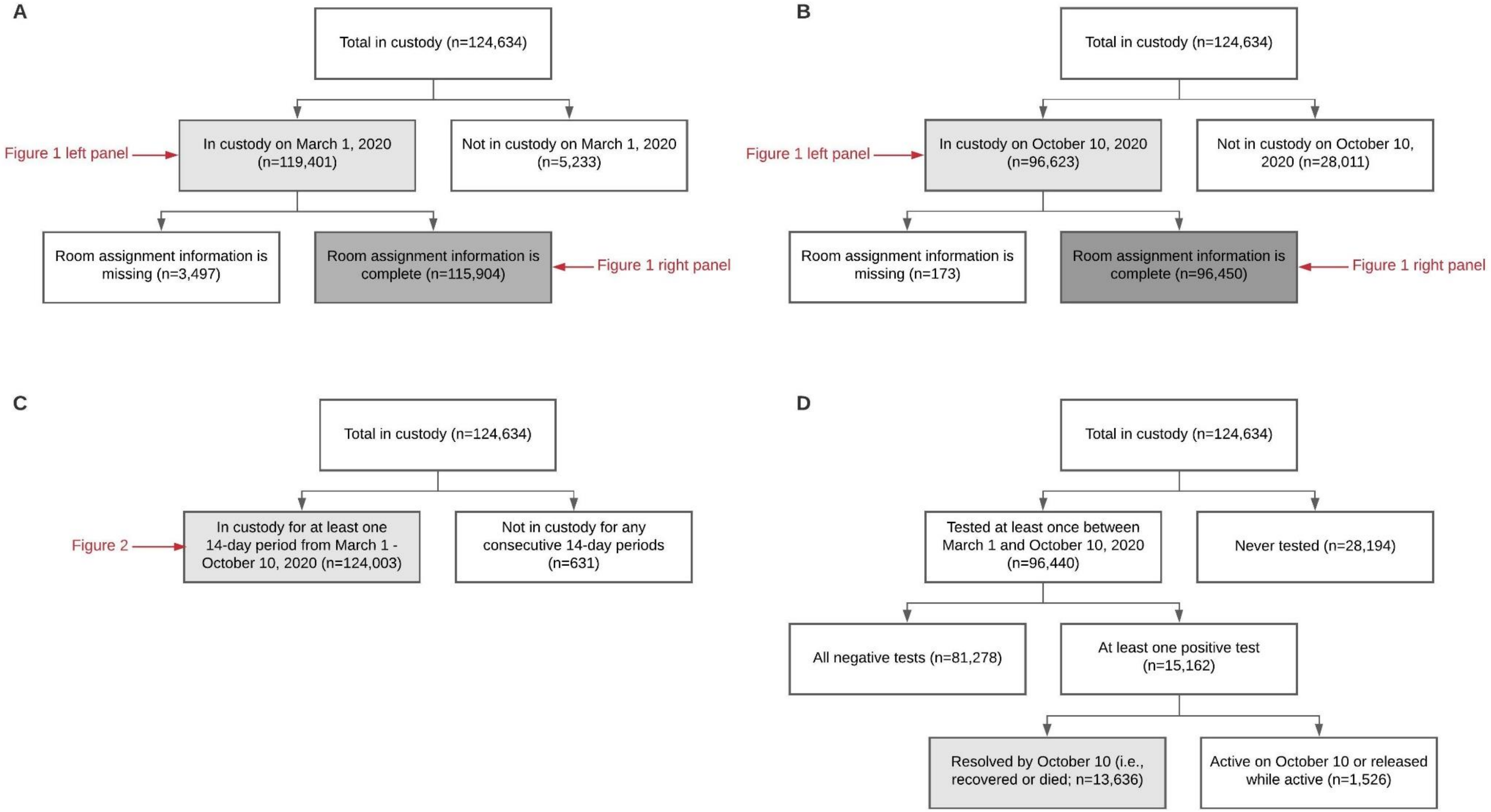


Most of the data displayed in Table 1 correspond to panel A and panel B (light grey box). The data on room occupancies in Table 1 correspond to the dark grey box in panels A and B. The data on labor participation in Table 1 corresponds to the light grey box in panel C. Finally, the data on Covid-19 outcomes (resolved cases, hospitalizations, intensive care unit admissions, deaths) correspond to the light grey box in panel D.

Figure S3: Across 35 CDCR prisons, we identified a set of persons who were in custody at any point in our study period (n=124,634). We identified a set of 19 prisons with ≥50 cumulative cases during the study period and ≥10 incident cases detected on at least one day in that period (n=71,942). We excluded individuals who were released prior, admitted after, or tested positive prior to the start of the outbreak (n=9,563). We excluded prisons with outbreaks seeded by mass introduction of cases (n=10,713; 3 prisons). Within prisons without mass introduction of cases (n=48,816; 16 prisons), we restricted our base analysis to prisons with outbreaks beginning on or before July 12, 2020 to allow for 90 follow-up days (n=26,760; 9 prisons), though in our sensitivity analyses, we varied this requirement. Finally, we included only persons who participated in testing during out study period (n=21,750). Of these, 743 (3.4%) arrived after March 1, 2020.

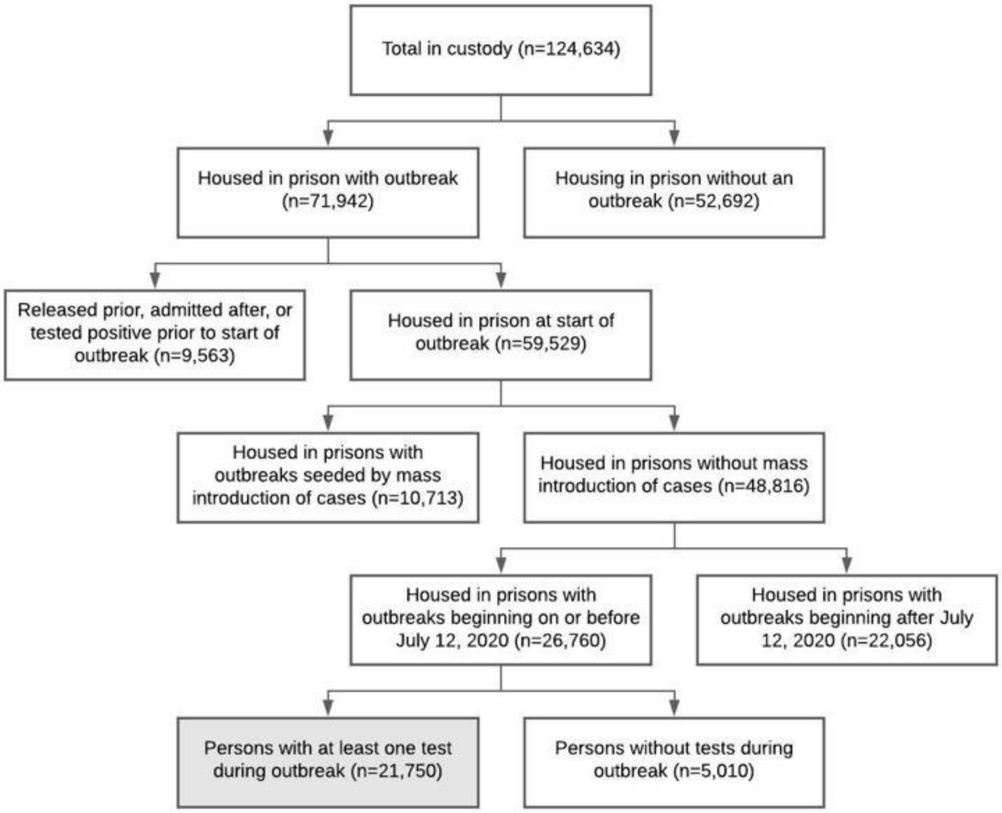

Figure S4: Testing across prisons with outbreaks by room type and labor status

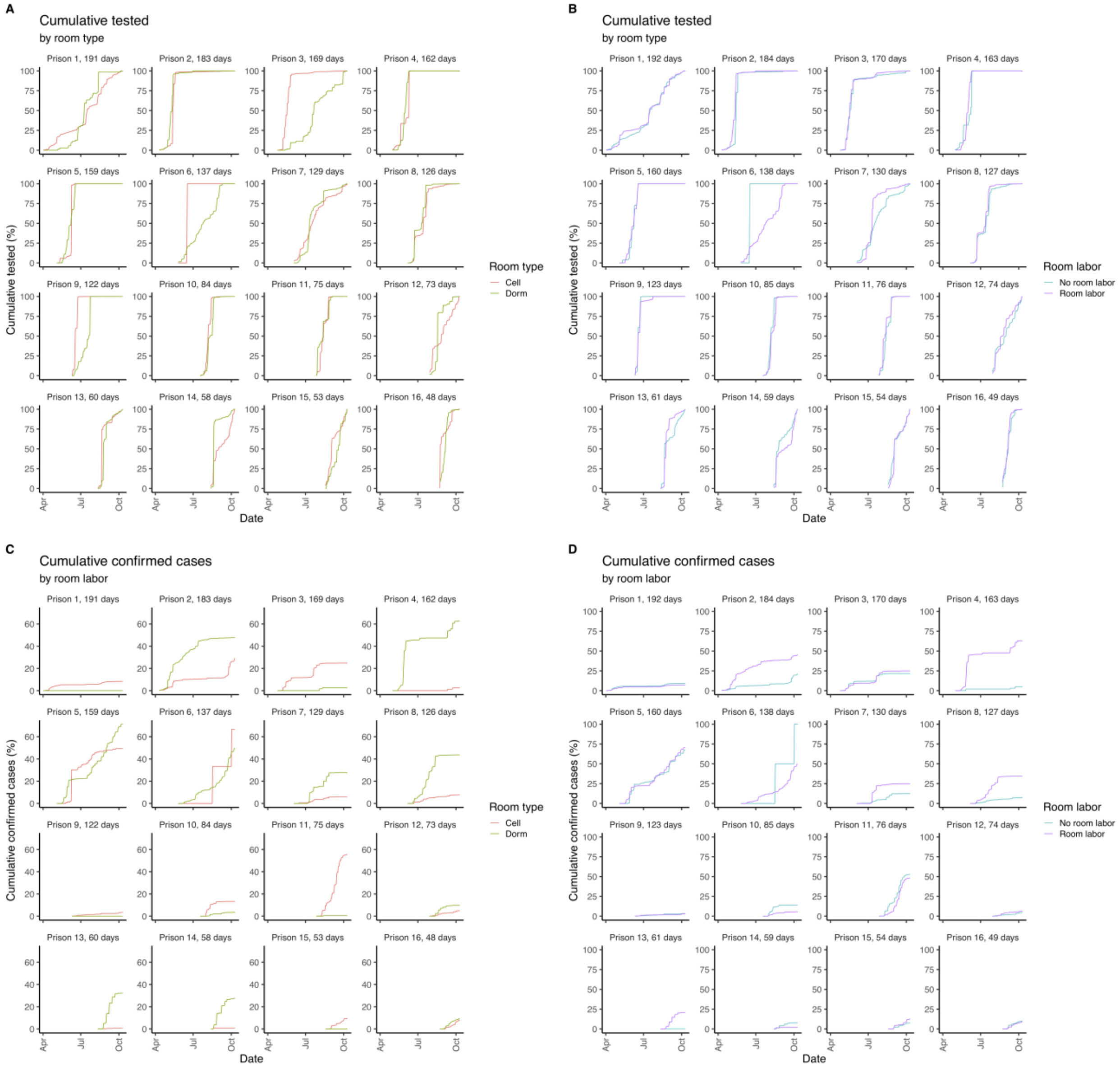


Notes: Graphs show the cumulative proportion of individuals tested (A & B) and confirmed positive (C & D) by room type and labor from the start of an outbreak, stratified by prison.

Figure S5: Room occupancy distributions by prison

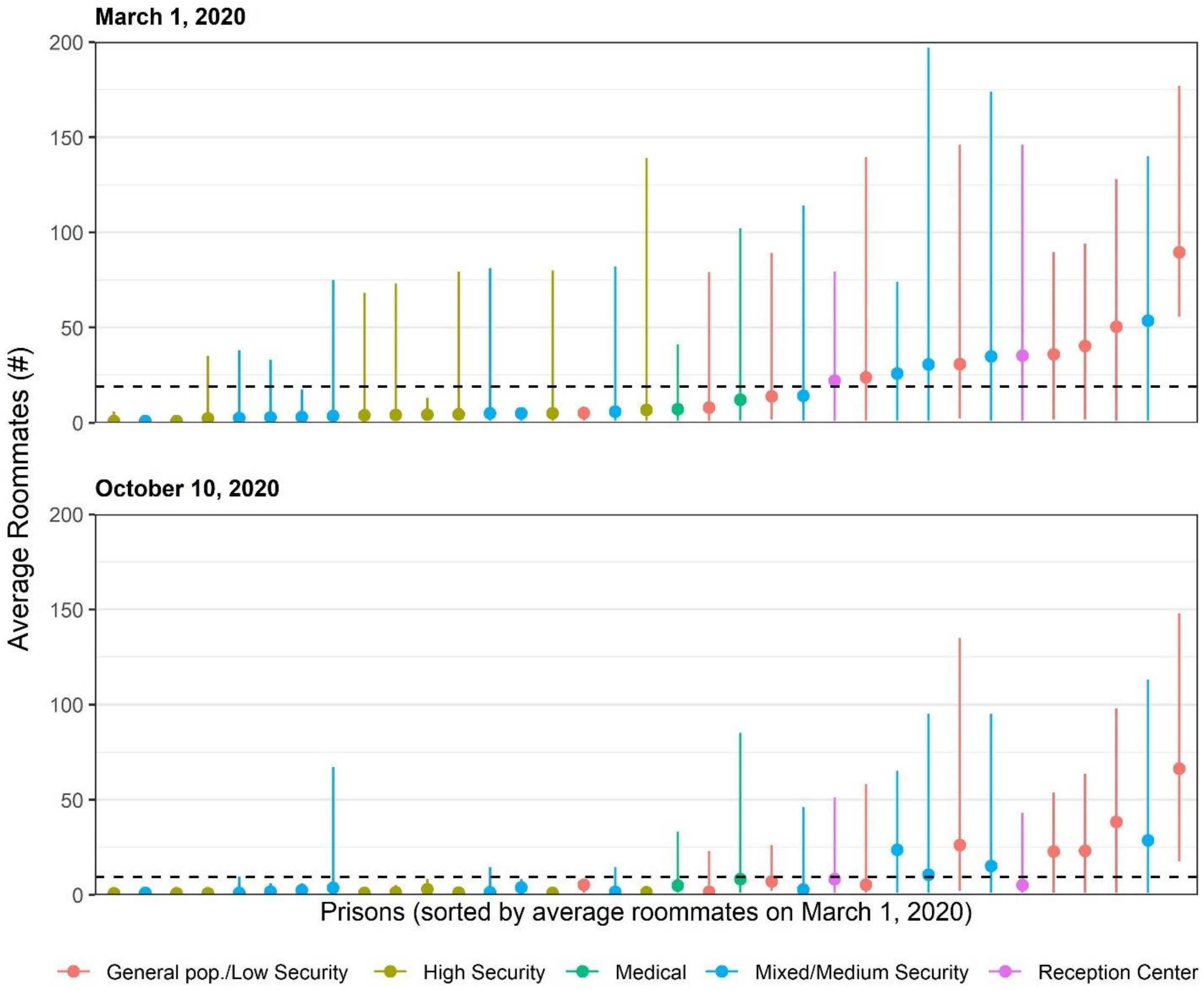


Note: Graph shows the number of roommates the average person in the prison system has, with 2.5th and 97.5th percentile numbers of roommates, by prison, on March 1, 2020 and October 10, 2020. Dashed lines indicate the average room occupancy on each date across all 35 prisons combined.

Figure S6: Room occupancy distributions by prison, excluding observed cases

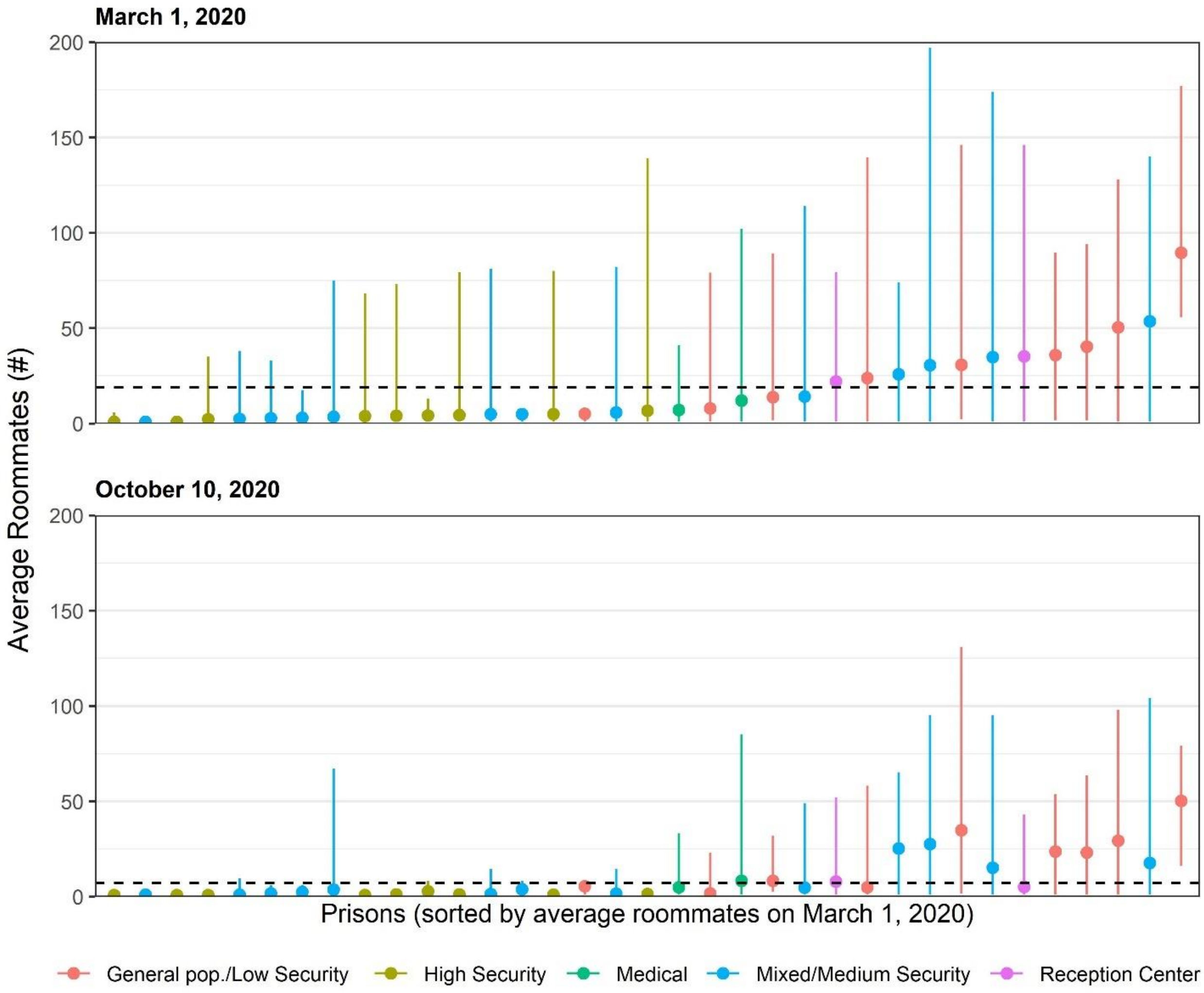


Note: Graph shows the number of roommates the average person in the prison system has, with 2.5th and 97.5th percentile numbers of roommates, by prison, on March 1, 2020 and October 10, 2020. Dashed lines indicate the average room occupancy on each date across all 35 prisons combined. Detected Covid-19 cases are excluded, so the graph shows presumed susceptible individuals.

Figure S7: Release & rehousing by age and security level

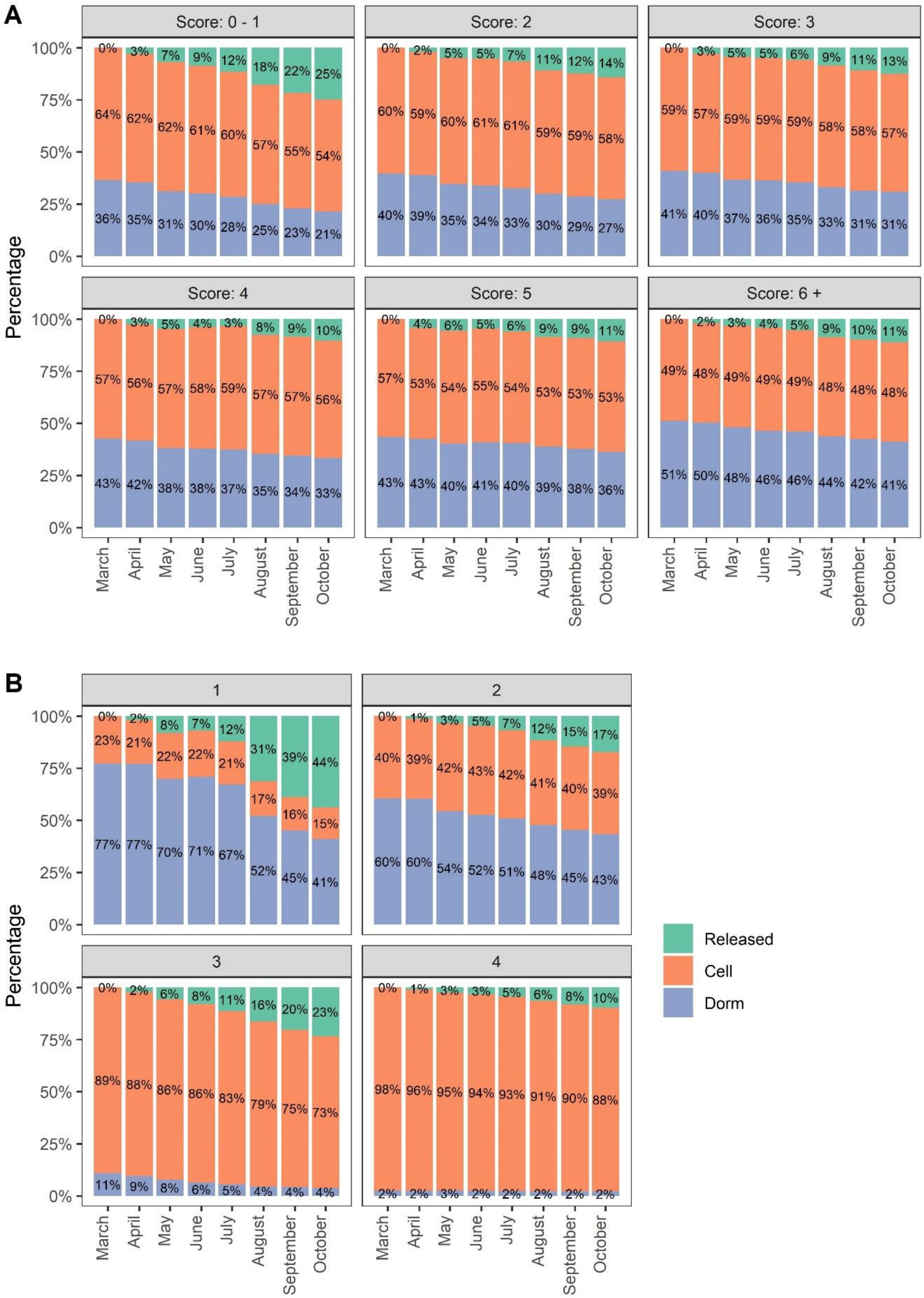

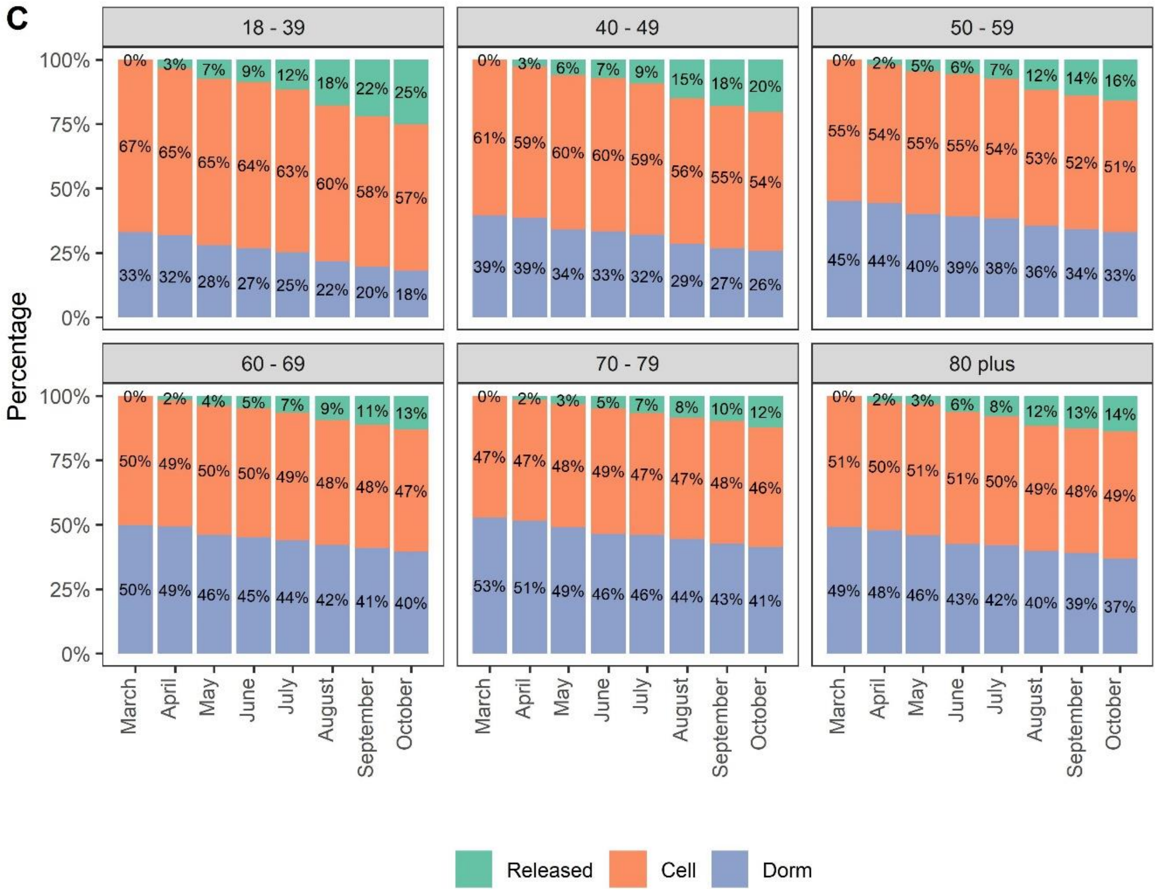


Notes: Graphs show the proportion of individuals in custody on March 1, 2020 that reside in cells, dorms, or have been released, stratified by (A) Covid-19 risk score, (B) security level, and (C) age. Data represent an individual's status at the first of each month and cover all prisons from March 1, 2020 through October 1, 2020.

Figure S8: Biweekly participation in labor and other out-of-room activities by Covid-19 risk score

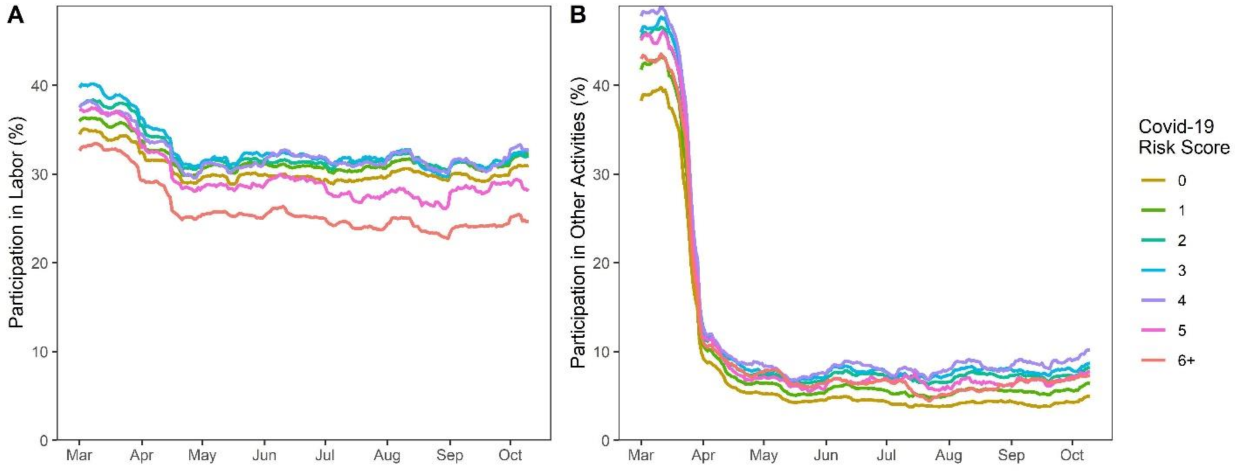


Graph shows rolling average participation in activities, defined as whether an individual participated in labor or other activities with at least one other person during any day in the past 2 weeks. Panels show (A) labor participation by Covid-19 risk score; (B) other participation by Covid-19 risk score. Data cover all prisons from March 1, 2020 through October 10, 2020.

Figure S9: Activity participation by age and prison type

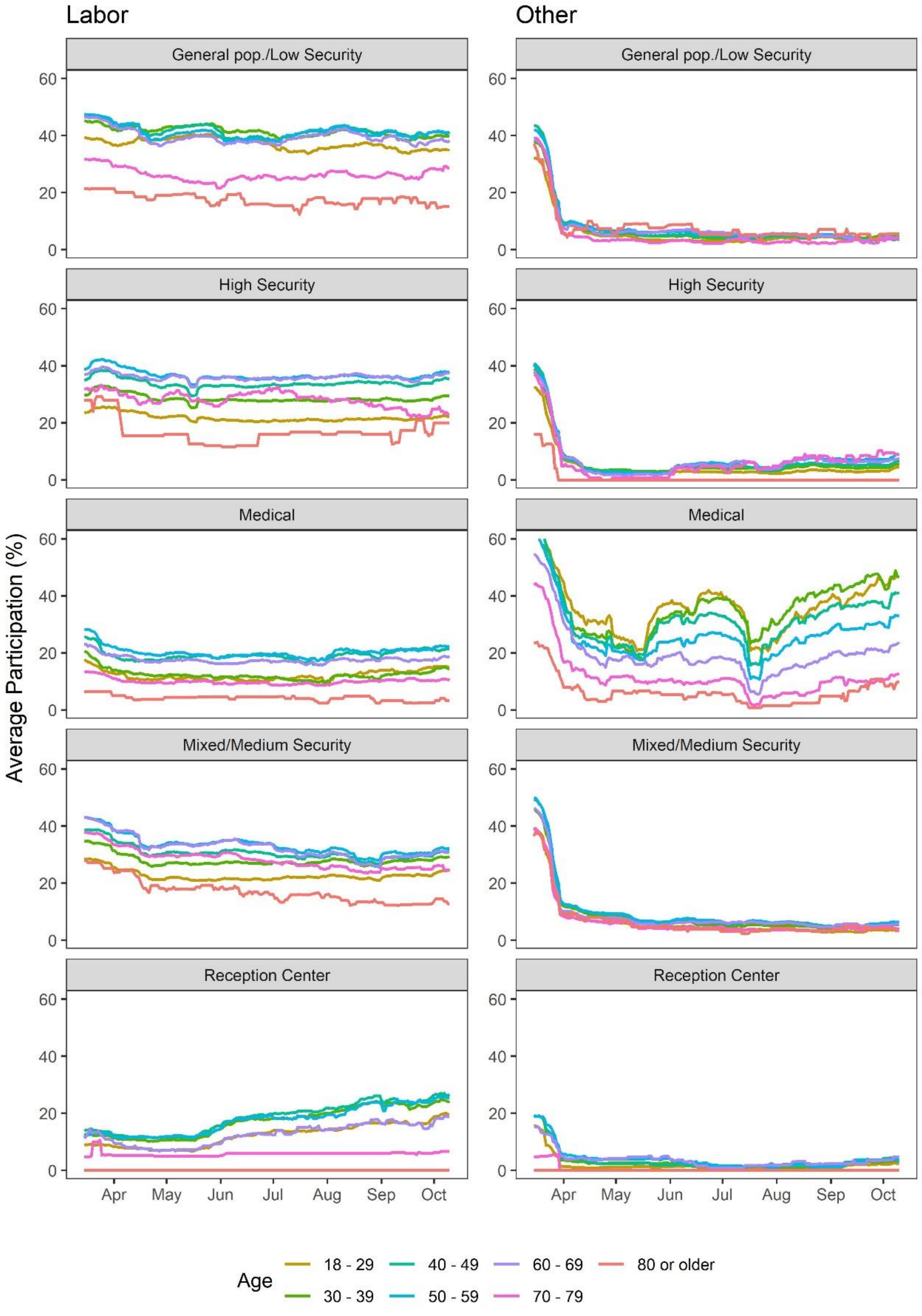


Notes: Graph shows rolling average participation in activities, defined as whether an individual participated in labor or other activities with at least one other person during any day in the past 2 weeks. Data cover all prisons from March 1, 2020 - October 10, 2020, stratified by prison type.

Figure S10: Activity participation by Covid-19 risk score and prison type

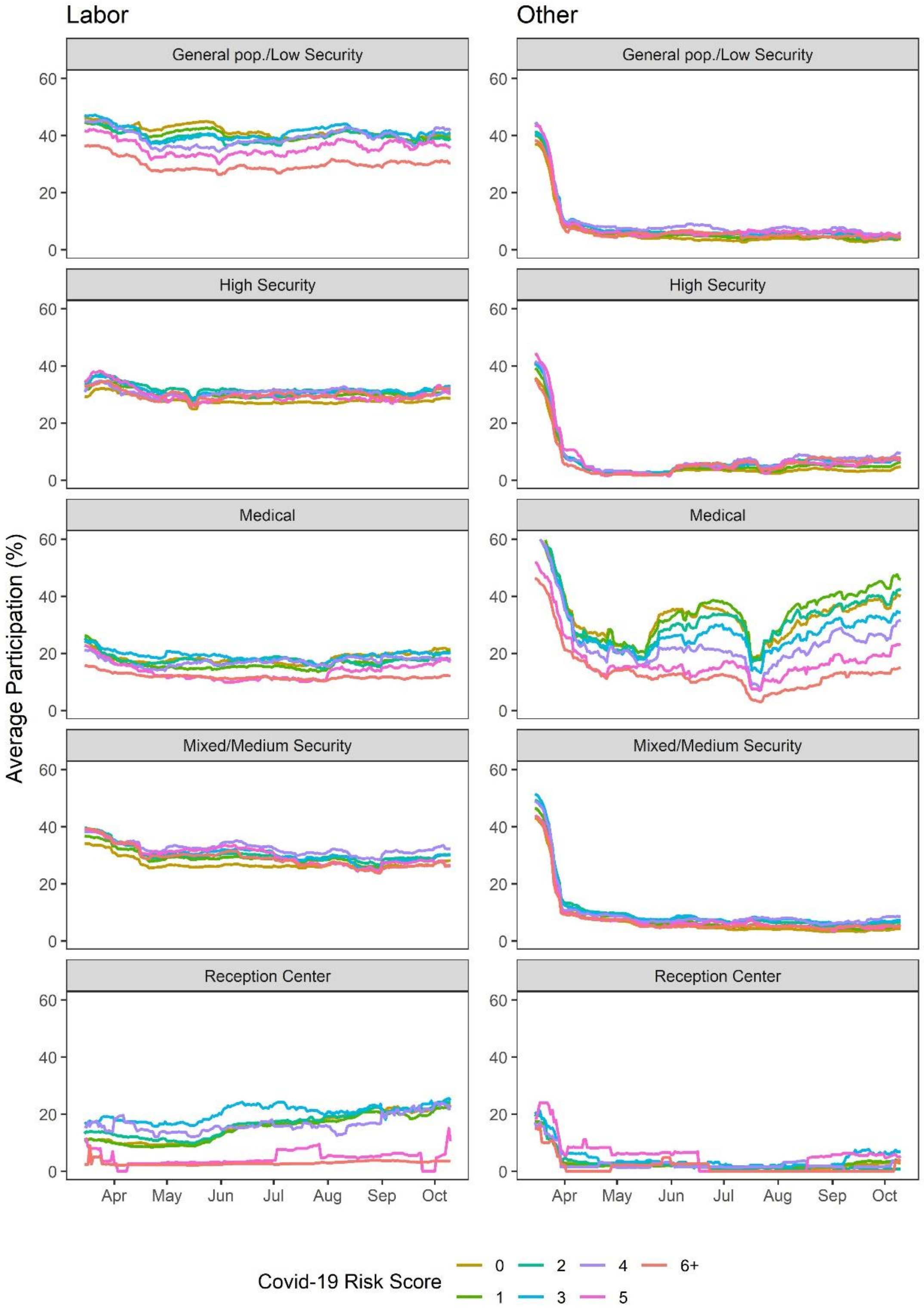


Notes: Graph shows rolling average participation in activities, defined as whether an individual participated in labor or other activities with at least one other person during any day in the past 2 weeks. Data cover all prisons from March 1, 2020 - October 10, 2020, stratified by prison type.

Figure S11: Release and rehousing by Covid-19 risk score in outbreak and non-outbreak prisons

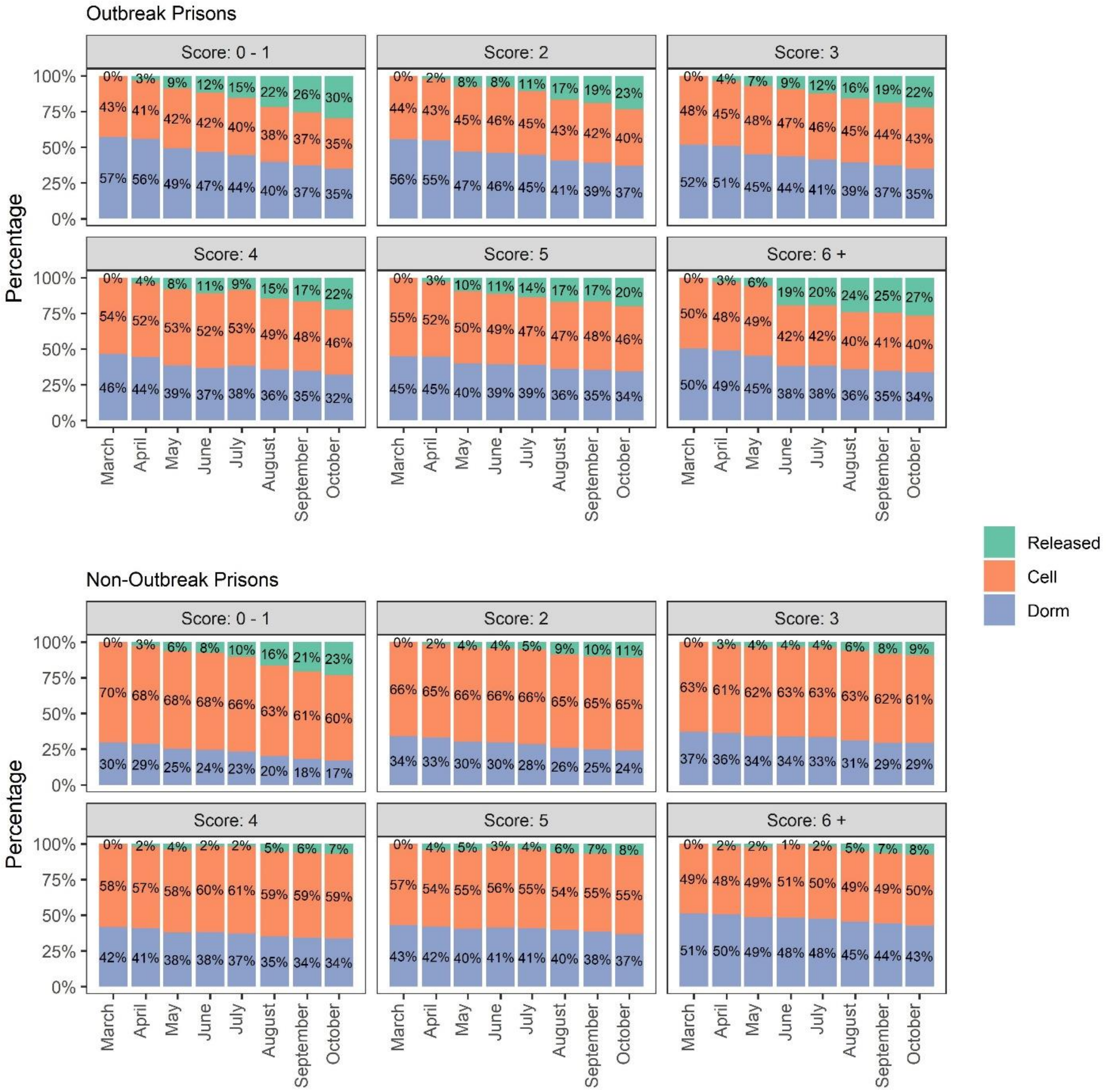


Notes: Graphs show the proportion of individuals in custody on March 1, 2020 that reside in cells, dorms, or have been released, stratified by Covid-19 risk score and whether they were housed in one of the nine outbreak prisons. Data represent an individual's status at the first of each month and only the nine outbreak prisons from March 1, 2020 through October 1, 2020.

Figure S12: Release and rehousing by security level in outbreak and non-outbreak prisons

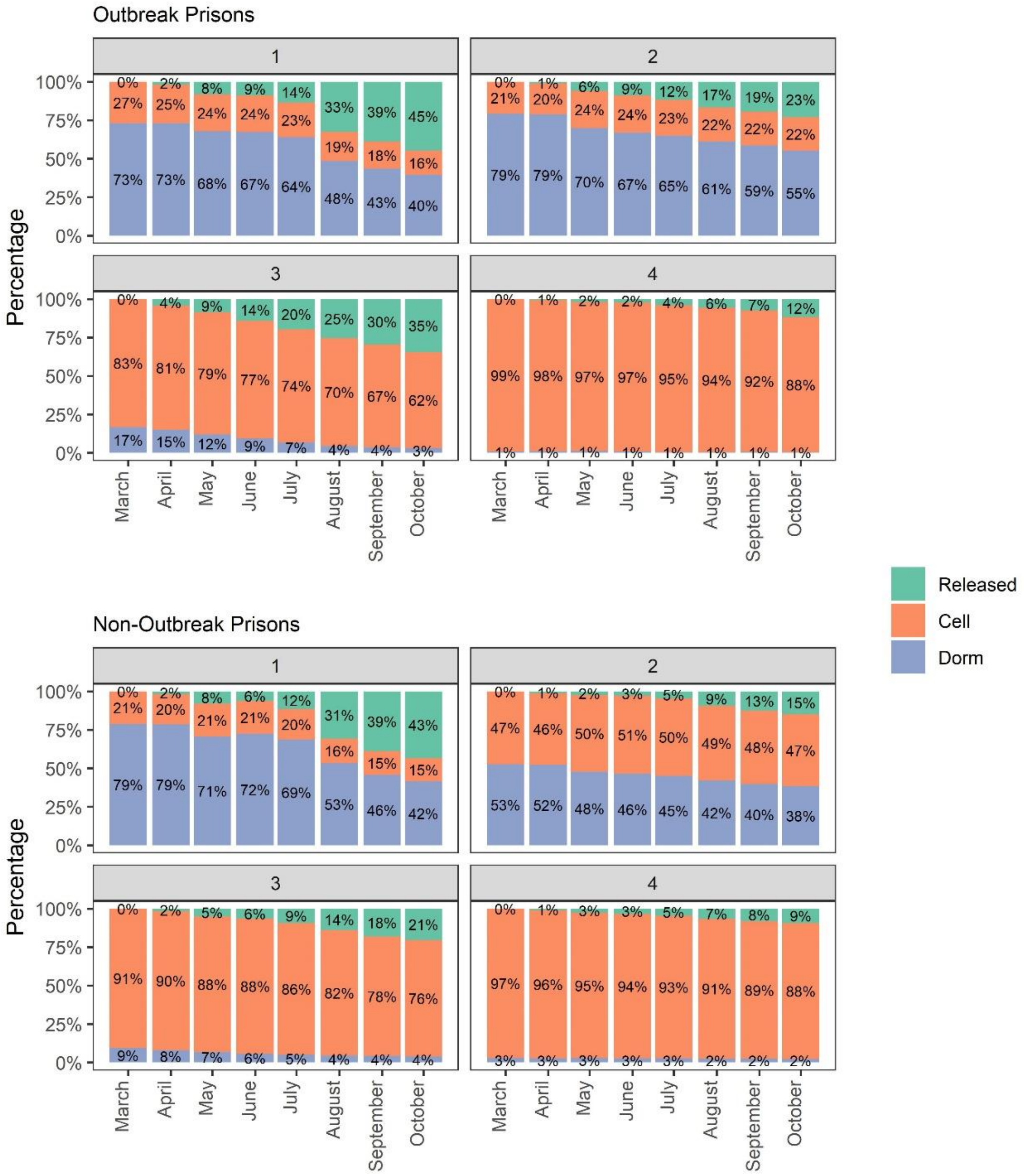


Notes: Graphs show the proportion of individuals in custody on March 1, 2020 that reside in cells, dorms, or have been released, stratified by security level and whether they were housed in one of the nine outbreak prisons. Data represent an individual's status at the first of each month and only the nine outbreak prisons from March 1, 2020 through October 1, 2020.

Figure S13: Release and rehousing by age in outbreak and non-outbreak prisons

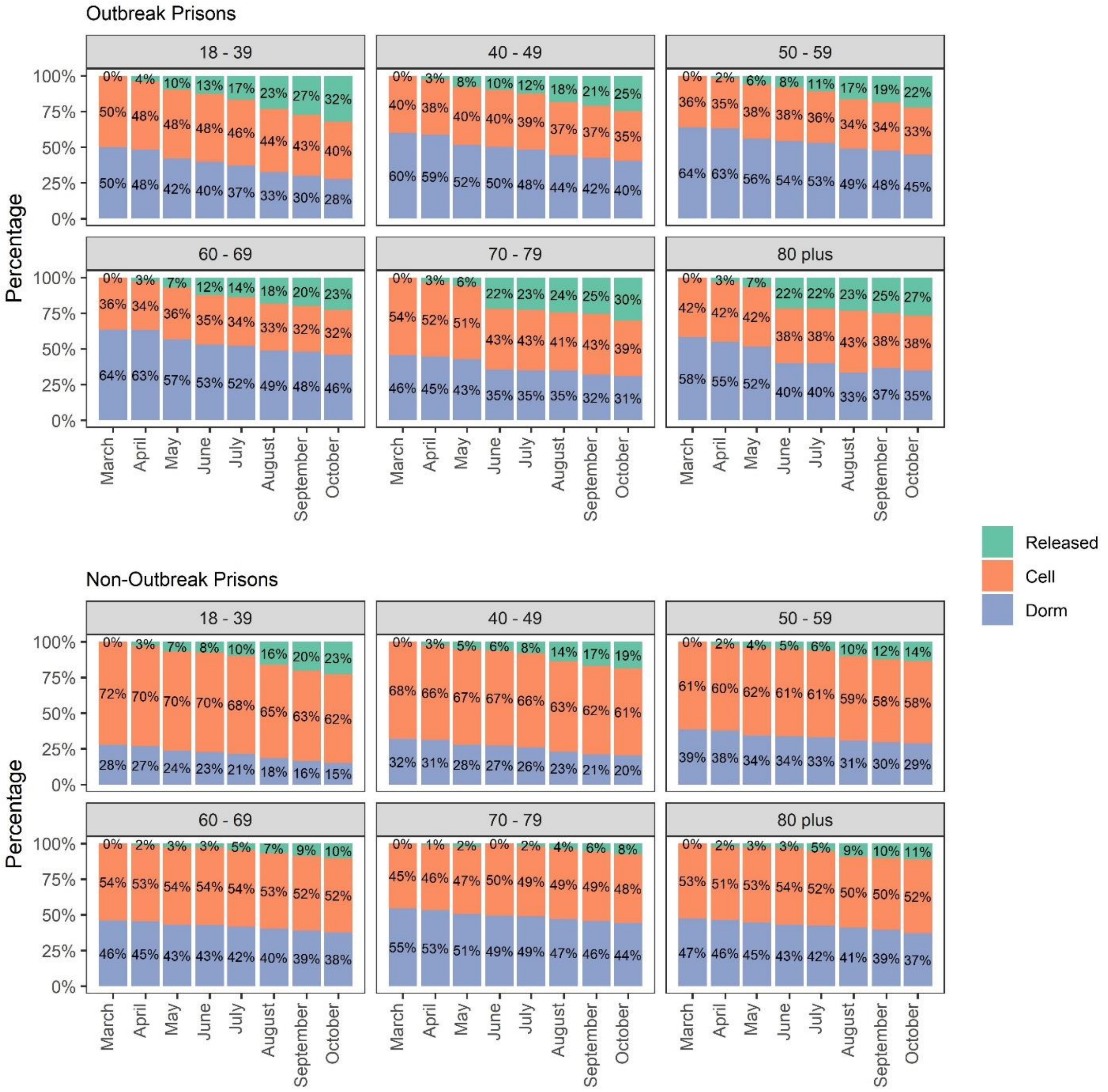


Notes: Graphs show the proportion of individuals in custody on March 1, 2020 that reside in cells, dorms, or have been released, stratified by age and whether they were housed in one of the nine outbreak prisons. Data represent an individual's status at the first of each month and only the nine outbreak prisons from March 1, 2020 through October 1, 2020.

Figure S14: Participation in labor and other activities in outbreak and non-outbreak prisons

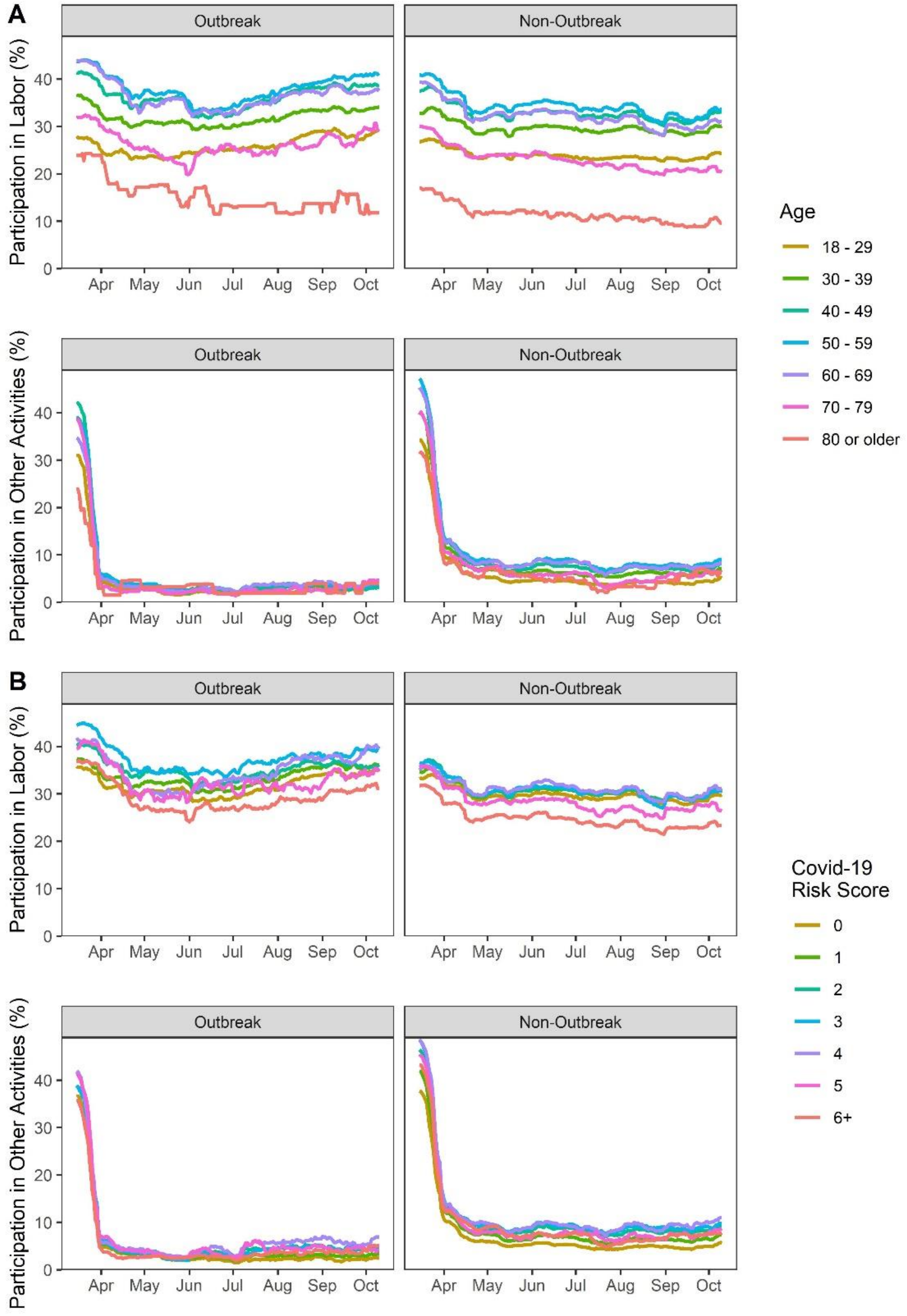


Notes: Graph shows rolling average participation in activities, defined as whether an individual participated in labor or other activities with at least one other person during any day in the past 2 weeks. Panels show (A) labor and other participation by age and (B) labor and other participation by Covid-19 risk score. Data cover only the nine outbreak prisons from March 1, 2020 through October 10, 2020.

Table S5: Change in prison characteristics between March and October among outbreak and non-outbreak prisons

| | Outbreak Prisons | | Non-Outbreak Prisons | |
|---|---|---|---|---|
| | March 1, 2020 (n=30,633) | October 10, 2020 (n=21,889) | March 1, 2020 (n=88,768) | October 10, 2020 (n=74,734) |
| **Demographic Characteristics** | | | | |
| Age, mean (SD) | 41.4 (12.5) | 42.1 (12.5) | 41.3 (13.1) | 41.8 (13.3) |
| 18-29 | 5853 (19.1%) | 3786 (17.3%) | 18497 (20.8%) | 15123 (20.2%) |
| 30-39 | 9417 (30.7%) | 6524 (29.8%) | 27156 (30.6%) | 22094 (29.6%) |
| 40-49 | 7323 (23.9%) | 5453 (24.9%) | 19254 (21.7%) | 16145 (21.6%) |
| 50-59 | 5154 (16.8%) | 3953 (18.1%) | 14124 (15.9%) | 12481 (16.7%) |
| 60-69 | 2234 (7.3%) | 1706 (7.8%) | 7543 (8.5%) | 6855 (9.2%) |
| 70-79 | 584 (1.9%) | 414 (1.9%) | 1886 (2.1%) | 1762 (2.4%) |
| 80+ | 68 (0.2%) | 53 (0.2%) | 308 (0.3%) | 274 (0.4%) |
| Sex | | | | |
| Female | 1634 (5.3%) | 1174 (5.4%) | 3162 (3.6%) | 2031 (2.7%) |
| Male | 28999 (94.7%) | 20715 (94.6%) | 85606 (96.4%) | 72703 (97.3%) |
| Race | | | | |
| Black | 6970 (22.8%) | 5090 (23.3%) | 26951 (30.4%) | 23381 (31.3%) |
| Hispanic | 15297 (49.9%) | 11041 (50.4%) | 37690 (42.5%) | 31957 (42.8%) |
| White | 6507 (21.2%) | 4336 (19.8%) | 18108 (20.4%) | 14287 (19.1%) |
| Other/Unknown | 1859 (6.1%) | 1422 (6.5%) | 6019 (6.8%) | 5109 (6.8%) |
| Security level | | | | |
| 1 | 3873 (12.6%) | 2116 (9.7%) | 8593 (9.7%) | 5224 (7.0%) |
| 2 | 16167 (52.8%) | 12535 (57.3%) | 36458 (41.1%) | 32699 (43.8%) |
| 3 | 4576 (14.9%) | 3009 (13.7%) | 16524 (18.6%) | 13586 (18.2%) |
| 4 | 4653 (15.2%) | 4099 (18.7%) | 25008 (28.2%) | 22989 (30.8%) |
| Number of roommates, mean (SD) | 34.5 (46.7) | 21.1 (35.4) | 13.9 (33.3) | 5.8 (14.7) |
| Cells | 12511 (40.8%) | 11204 (51.2%) | 59062 (66.5%) | 55422 (74.2%) |
| Dorms | 15767 (51.5%) | 10562 (48.3%) | 28564 (32.2%) | 19262 (25.8%) |
| Work | | | | |
| Worked previous two weeks | 11262 (38.0%) | 7755 (35.9%) | 30685 (34.9%) | 22183 (29.9%) |
| **Clinical** | | | | |
| Covid-19 risk score, mean (SD) | 1.3 (1.8) | 1.3 (1.8) | 1.4 (2.0) | 1.5 (2.1) |
| Covid-19 risk score of 3+ | 4558 (14.9%) | 3479 (15.9%) | 15262 (17.2%) | 14228 (19.0%) |
| Pre-existing conditions | | | | |
| Any pre-existing condition* | 10736 (35.0%) | 8229 (37.6%) | 33577 (37.8%) | 30289 (40.5%) |
| Advanced liver disease | 955 (3.1%) | 630 (2.9%) | 3282 (3.7%) | 2632 (3.5%) |
| Asthma | 3277 (10.7%) | 2444 (11.2%) | 11279 (12.7%) | 9941 (13.3%) |
| Cancer | 725 (2.4%) | 581 (2.7%) | 2302 (2.6%) | 2125 (2.8%) |
| COPD | 577 (1.9%) | 442 (2.0%) | 2497 (2.8%) | 2264 (3.0%) |
| CVD | 1109 (3.6%) | 770 (3.5%) | 4345 (4.9%) | 3643 (4.9%) |
| Diabetes | 2075 (6.8%) | 1572 (7.2%) | 6491 (7.3%) | 5976 (8.0%) |

| | | | | |
|---|---|---|---|---|
| HIV | 316 (1.0%) | 193 (0.9%) | 640 (0.7%) | 531 (0.7%) |
| Hypertension | 6001 (19.6%) | 4848 (22.1%) | 19709 (22.2%) | 18476 (24.7%) |
| Immunocompromised | 320 (1.0%) | 246 (1.1%) | 1057 (1.2%) | 989 (1.3%) |
| Body Mass Index, mean (SD) | 28.9 (5.5) | 29.0 (5.5) | 28.6 (5.3) | 28.9 (5.5) |
| Body Mass Index > 40 | 1031 (3.4%) | 748 (3.4%) | 2911 (3.3%) | 2775 (3.7%) |
| **Covid-19 testing and cases (Cumulative persons since January 1, 2020)** | | | | |
| Individuals Tested | | 25415 | | 71025 |
| Positive (% of Tested) | | 8950 (35.2%) | | 6212 (8.7%) |
| Resolved Positive (% of Tested) | | 7871 (31.0%) | | 5765 (8.1%) |
| Hospitalized (% of Resolved) | | 268 (3.4%) | | 174 (3.0%) |
| ICU (% of Resolved) | | 12 (0.2%) | | 25 (0.4%) |
| Deaths (% of Resolved) | | 36 (0.5%) | | 32 (0.6%) |

COPD = Chronic Obstructive Pulmonary Disease; CVD = Cardiovascular Disease; HIV = Human Immunodeficiency Virus; ICU = Intensive Care Unit; SD = Standard Deviation.

Figure S15. Tests and confirmed cases in outbreak prisons

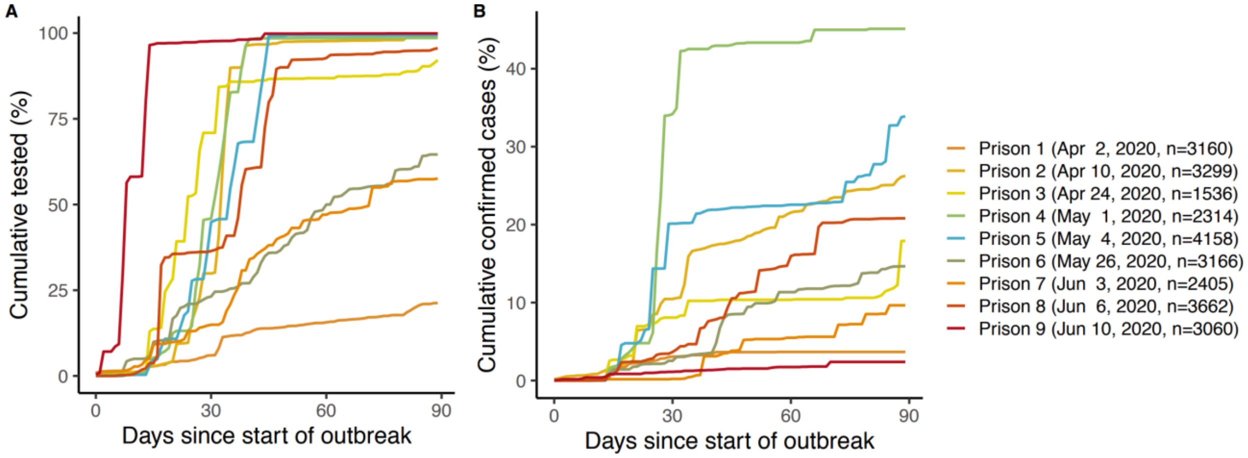


Graphs show the distribution of (A) cumulative individuals tested and (B) cumulative confirmed cases across outbreak prisons. Data includes 90 days follow-up for 9 prisons.

Figure S16: Events by day of outbreak

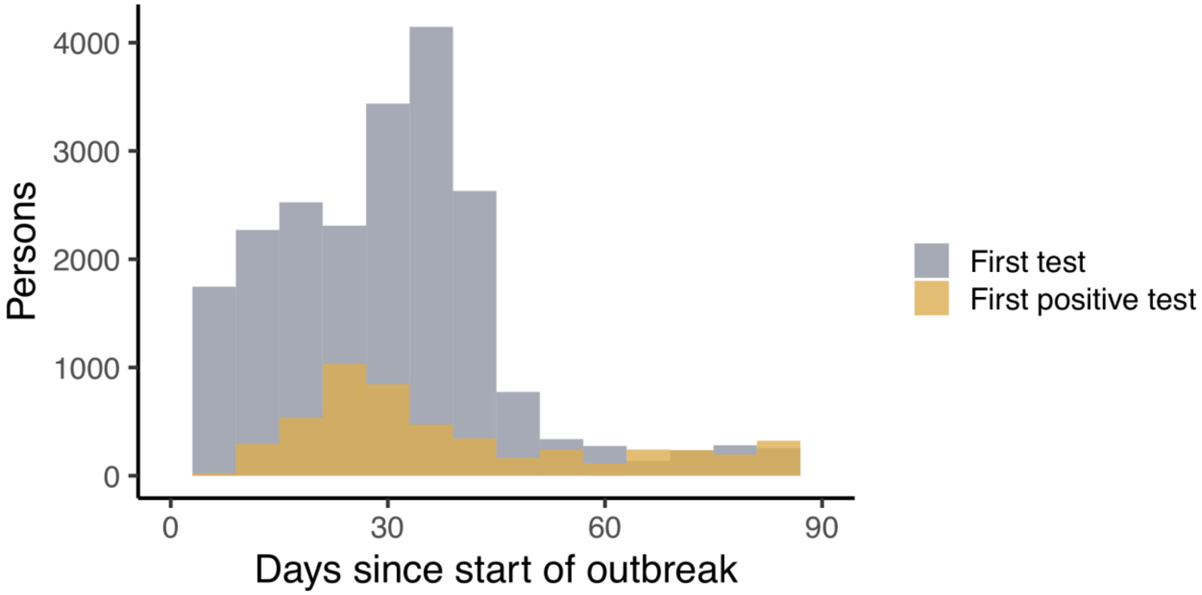


Notes: Graph shows the distribution of an individual's time to first test and time to first positive test (days since start of outbreak). Data include 90 days follow-up for 9 prisons.

Figure S17: Proportionality assumption

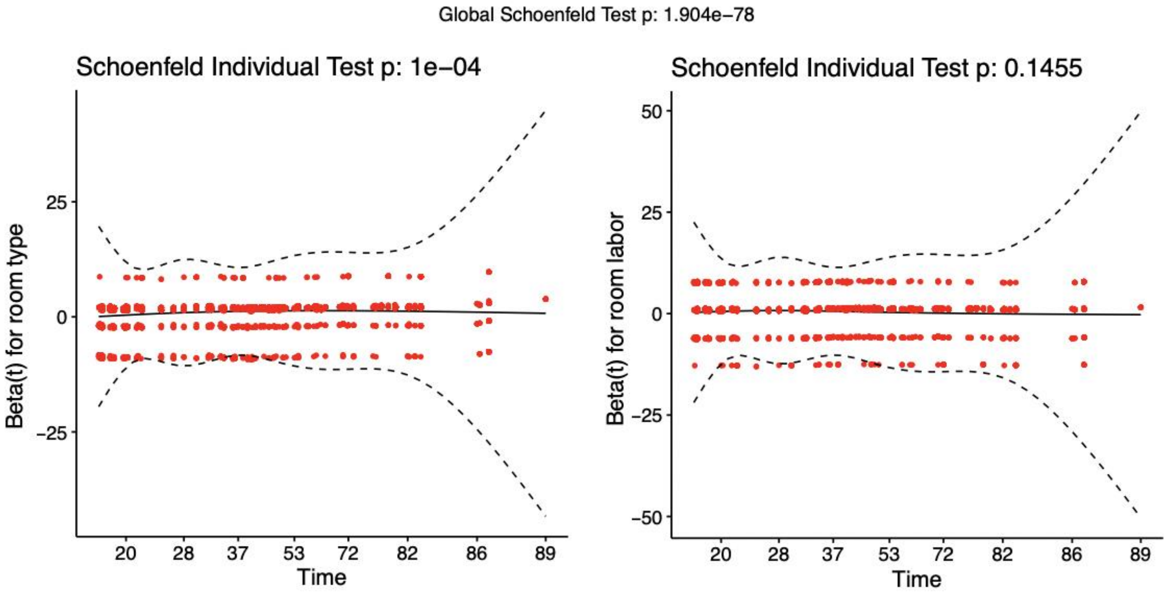


Note: Visual inspection of the Schoenfeld residuals supports the validity of the proportional hazard assumptions in the Cox regression models.

Table S6: Survival analysis - sensitivity analysis results

| * p < 0.05<br>** p < 0.01<br>*** p < 0.001 | # Prisons | # Rooms | # Persons | **Censor by last test date**<br>Adjusted Hazard Ratio | | **Censor by last date in outbreak prison**<br>Adjusted Hazard Ratio | |
|---|---|---|---|---|---|---|---|
| | | | | **Room type (Dorm)** | **Labor (Room labor)** | **Room type (Dorm)** | **Labor (Room labor)** |
| **Base case (90 days to follow up)** | 9 | 6,928 | 21,750 | 2.51 ***<br>(2.25 – 2.80) | 1.56 ***<br>(1.39 – 1.74) | 2.99 ***<br>(2.68 – 3.34) | 1.40 ***<br>(1.26 – 1.57) |
| **Removal of prisons with differences in testing rates for cells vs dorms**[x] | 8 | 6,060 | 20,335 | 2.67 ***<br>(2.37 – 3.01) | 1.72 ***<br>(1.51 – 1.96) | 2.67 ***<br>(2.37 – 3.01) | 1.47 ***<br>(1.30 – 1.67) |
| **Adding CDCR Covid-19 risk score**[xx] | 9 | 6,928 | 21,750 | 2.50 ***<br>(2.24 – 2.79) | 1.55 ***<br>(1.39 – 1.73) | 2.98 ***<br>(2.67 – 3.33) | 1.39 ***<br>(1.24 – 1.55) |
| **Adding Security level**[xxx] | 9 | 6,921 | 21,732 | 2.46 ***<br>(2.16 – 2.80) | 1.58 ***<br>(1.41 – 1.77) | 3.10 ***<br>(2.72 – 3.52) | 1.46 ***<br>(1.30 – 1.63) |
| **Adding Age, Sex, Race/Ethnicity, Pre-existing conditions**[xxxx] | 9 | 6,928 | 21,750 | 2.49 ***<br>(2.23 – 2.78) | 1.56 ***<br>(1.39 – 1.74) | 2.96 ***<br>(2.65 – 3.31) | 1.40 ***<br>(1.25 – 1.56) |
| **60 days to follow up** | 13 | 12,417 | 31,009 | 1.58 ***<br>(1.45 – 1.72) | 1.13 **<br>(1.04 – 1.22) | 1.75 ***<br>(1.61 – 1.91) | 1.12 **<br>(1.03 – 1.21) |
| **120 days to follow up** | 9 | 7,519 | 23,648 | 2.84 ***<br>(2.56 – 3.14) | 1.60 ***<br>(1.45 – 1.78) | 2.81 ***<br>(2.54 – 3.11) | 1.42 ***<br>(1.28 – 1.57) |
| **150 days to follow up** | 5 | 4,219 | 13,016 | 2.16 ***<br>(1.90 – 2.44) | 1.35 ***<br>(1.20 – 1.51) | 2.27 ***<br>(2.01 – 2.57) | 1.26 ***<br>(1.13 – 1.41) |
| **No follow up censoring** | 16 | 16,342 | 40,369 | 2.03 ***<br>(1.90 – 2.17) | 1.15 ***<br>(1.08 – 1.23) | 1.66 ***<br>(1.55 – 1.78) | 1.14 ***<br>(1.07 – 1.22) |

[x] Excluding prison 3: based on large differences in testing rates for cells vs dorms at the start of the outbreak.

[xx] Adding CDCR Covid-19 risk score to the model: The CDCR Covid-19 risk score was used as a proxy variable to control for differences in testing priority according to medical vulnerabilities. We top-coded CDCR Covid-19 risk to 6. We used the model Surv(time, pos) ~ prison + room type + labor + Covid-19 risk.

[xxx] Adding Security level to the model: Security level was used as a proxy variable to control for differences in testing priority according to medical vulnerabilities. We used the model Surv(time, pos) ~ prison + room type + labor + security level.

[xxxx] Adding Age, Sex, Race/Ethnicity, Pre-existing conditions to the model: Age, Sex, Race/Ethnicity, and several pre-existing conditions were used as controls. We used the model Surv(time, pos) ~ prison + room type + labor + advanced liver disease + asthma + cancer + chronic obstructive pulmonary disease + cardiovascular disease + diabetes + HIV + hypertension + immunocompromised.

Table S7: Standard errors -- unclustered & clustered by prison and room

| | | | Unclustered | | | Marginal: Clustered by prison | | | Marginal: Clustered by room | | |
|---|---|---|---|---|---|---|---|---|---|---|---|
| | coef | exp(coef) | se | z | p | se | z | p | se | z | p |
| Dorm | 0.92 | 2.51 | 0.06 | 16.6 | < 2e-16 | 0.15 | 6.13 | 8.71E-10 | 0.32 | 2.84 | 0.00452 |
| Room labor | 0.44 | 1.56 | 0.06 | 7.80 | 5.98E-15 | 0.12 | 3.80 | 1.45 E-4 | 0.15 | 2.97 | 0.00298 |

Notes: Table shows standard errors for unclustered, clustered by prison, and clustered by room models. Data include 90 days follow-up for 9 prisons.

Table S8: Regression coefficients

| | | **Base case (90 days to follow up)** | **Removal of prisons with differences in testing rates** | **Adding Covid-19 risk score** | **Adding security level** | **Adding Age, Sex, Race/Ethnicity, Pre-existing conditions** | **60 days to follow up** | **120 days to follow up** | **150 days to follow up** | **All outbreaks** |
|---|---|---|---|---|---|---|---|---|---|---|
| *Predictors* | | *Adjusted odds ratio* | *Adjusted odds ratio* | *Adjusted odds ratio* | *Adjusted odds ratio* | *Adjusted odds ratio* | *Adjusted odds ratio* | *Adjusted odds ratio* | *Adjusted odds ratio* | *Adjusted odds ratio* |
| Room type | Cell (Ref) | 1.00 | 1.00 | 1.00 | 1.00 | 1.00 | 1.00 | 1.00 | 1.00 | 1.00 |
| | Dorm | 2.51 *** (2.25 – 2.80) | 2.67 *** (2.37 – 3.01) | 2.50 *** (2.25 – 2.79) | 2.71 *** (2.40 – 3.06) | 2.49 *** (2.23 – 2.78) | 1.58 *** (1.45 – 1.72) | 2.84 *** (2.56 – 3.14) | 2.16 *** (1.90 – 2.44) | 2.03 *** (1.90 – 2.17) |
| Room labor | None (Ref) | 1.00 | 1.00 | 1.00 | 1.00 | 1.00 | 1.00 | 1.00 | 1.00 | 1.00 |
| | Any | 1.56 *** (1.39 – 1.74) | 1.72 *** (1.51 – 1.96) | 1.55 *** (1.39 – 1.74) | 1.58 *** (1.41 – 1.77) | 1.56 *** (1.39 – 1.74) | 1.13 ** (1.04 – 1.22) | 1.60 *** (1.45 – 1.78) | 1.35 *** (1.20 – 1.51) | 1.15 *** (1.08 – 1.23) |
| Covid-19 risk score | (continuous) | | | 1.02 * (1.00 – 1.04) | | | | | | |
| Security level | (continuous) | | | | 1.07 ** (1.02 – 1.13) | 1.07 * (1.02 – 1.13) | | | | |
| Age | (continuous) | | | | | 1.00 (1.00 – 1.00) | | | | |
| Sex | Female (Ref) | | | | | 1.00 | | | | |
| | Male | | | | | 0.92 (0.79 – 1.08) | | | | |
| Race | White (Ref) | | | | | 1.00 | | | | |
| | Black | | | | | 0.89 * (0.81 – 0.97) | | | | |
| | Hispanic | | | | | 1.02 (0.95 – 1.09) | | | | |
| | Other/Unknown | | | | | 0.89 (0.79 – 1.00) | | | | |
| Pre-existing conditions (Yes) | Advanced liver disease | | | | | 0.89 (0.75 – 1.06) | | | | |
| | Asthma | | | | | 1.04 (0.95 – 1.14) | | | | |
| | Cancer | | | | | 0.89 (0.75 – 1.05) | | | | |
| | COPD | | | | | 0.95 (0.80 – 1.13) | | | | |
| | CVD | | | | | 1.02 (0.89 – 1.18) | | | | |
| | Diabetes | | | | | 0.98 | | | | |

| | | | | | | | | | | |
|---|---|---|---|---|---|---|---|---|---|---|
| | | | | | | (0.88 – 1.09) | | | | |
| | HIV | | | | | 1.06<br>(0.82 – 1.36) | | | | |
| | Hypertension | | | | | 1.16 ***<br>(1.08 – 1.24) | | | | |
| | Immunocompromised | | | | | 1.00<br>(0.77 – 1.31) | | | | |
| | CCI | 0.81 ***<br>(0.74 – 0.89) | 0.84 ***<br>(0.76 – 0.92) | 0.81 ***<br>(0.73 – 0.88) | 0.82 ***<br>(0.74 – 0.90) | 0.83 ***<br>(0.76 – 0.92) | 0.74 ***<br>(0.66 – 0.83) | 0.69 ***<br>(0.63 – 0.76) | | 0.74 ***<br>(0.68 – 0.81) |
| | CIM | 0.96<br>(0.88 – 1.05) | 0.98<br>(0.90 – 1.06) | 0.92<br>(0.83 – 1.01) | 0.99<br>(0.91 – 1.08) | 0.99<br>(0.89 – 1.09) | 1.15 **<br>(1.04 – 1.28) | 0.82 ***<br>(0.76 – 0.88) | 0.90 *<br>(0.84 – 0.98) | 0.64 ***<br>(0.60 – 0.69) |
| | CIW | 0.96<br>(0.83 – 1.12) | | 0.95<br>(0.81 – 1.10) | 1.05<br>(0.89 – 1.23) | | 1.15<br>(0.96 – 1.38) | 0.96<br>(0.84 – 1.11) | 0.91<br>(0.79 – 1.06) | 0.62 ***<br>(0.55 – 0.71) |
| | CMC | | | | | | 0.43 ***<br>(0.37 – 0.49) | | | 0.54 ***<br>(0.48 – 0.61) |
| | CRC | 0.53 ***<br>(0.47 – 0.58) | 0.51 ***<br>(0.46 – 0.57) | 0.52 ***<br>(0.47 – 0.58) | 0.53 ***<br>(0.47 – 0.59) | 0.55 ***<br>(0.49 – 0.61) | 0.67 ***<br>(0.59 – 0.76) | 0.39 ***<br>(0.36 – 0.42) | | 0.76 ***<br>(0.72 – 0.81) |
| | CTF | | | | | | 0.21 ***<br>(0.18 – 0.26) | | | 0.33 ***<br>(0.28 – 0.39) |
| | CVSP | 1.73 ***<br>(1.60 – 1.88) | 1.74 ***<br>(1.61 – 1.89) | 1.72 ***<br>(1.59 – 1.87) | 1.74 ***<br>(1.60 – 1.88) | 1.75 ***<br>(1.61 – 1.90) | 1.69 ***<br>(1.55 – 1.85) | 1.53 ***<br>(1.41 – 1.65) | 1.44 ***<br>(1.35 – 1.54) | 1.01<br>(0.95 – 1.08) |
| | FSP | | | | | | 1.46 ***<br>(1.31 – 1.62) | | | 3.35 ***<br>(3.08 – 3.65) |
| | ISP | 0.57 ***<br>(0.45 – 0.73) | 0.61 ***<br>(0.48 – 0.78) | 0.57 ***<br>(0.45 – 0.73) | 0.60 ***<br>(0.46 – 0.76) | 0.61 ***<br>(0.48 – 0.79) | 0.45 ***<br>(0.34 – 0.60) | 0.16 ***<br>(0.13 – 0.19) | | 0.19 ***<br>(0.15 – 0.23) |
| | LAC | 1.40 **<br>(1.14 – 1.72) | 1.51 ***<br>(1.23 – 1.86) | 1.36 **<br>(1.10 – 1.67) | 1.39 **<br>(1.13 – 1.71) | 1.45 ***<br>(1.17 – 1.79) | 1.89 ***<br>(1.54 – 2.33) | 0.43 ***<br>(0.35 – 0.52) | 0.26 ***<br>(0.22 – 0.31) | 0.13 ***<br>(0.11 – 0.16) |
| | SATF | | | | | | 1.10<br>(0.97 – 1.25) | | | 1.61 ***<br>(1.43 – 1.81) |
| | WSP | 0.89<br>(0.76 – 1.04) | 0.97<br>(0.83 – 1.14) | 0.89<br>(0.76 – 1.04) | 0.94<br>(0.80 – 1.10) | 0.96<br>(0.82 – 1.12) | 0.45 ***<br>(0.37 – 0.54) | 0.68 ***<br>(0.58 – 0.79) | | 0.61 ***<br>(0.53 – 0.70) |
| | NKSP | | | | | | | | | 0.67 **<br>(0.51 – 0.88) |
| | SVSP | | | | | | | | | 1.09<br>(0.89 – 1.33) |
| Prisons | VSP | | | | | | | | | 0.47 ***<br>(0.41 – 0.54) |
| Observations | | 21750 | 20335 | 21750 | 21732 | 21750 | 31009 | 23648 | 13016 | 40369 |

* *p<0.05* ** *p<0.01* *** *p<0.001*

Notes: Table shows the regression coefficients for all models in the sensitivity analyses. For analyses that included Covid-19 risk score as a covariate, we examined timing of testing and numbers of tests. Days from start of outbreak until first test had median of 25 days (mean: 32.4; IQR: 14-40 days) for those with Covid-19 risks scores <3 and a median of 25 days (mean: 29.7; IQR: 14-33 days) for those with Covid-19 risk scores ≥3. The cumulative of tests from start of outbreak among those who never tested positive was a median of 2 test (mean: 2.4; IQR: 1-3 tests) for those with Covid-19 risks scores <3 and a median of 2 test (mean: 3.0; IQR: 1-4 tests) for those with Covid-19 risk scores ≥3.
